# Positive association of abdominal subcutaneous fat with cardiometabolic risk in thin, young Indians: A window on the developmental trajectory of adiposity-based chronic disease (ABCD)?

**DOI:** 10.64898/2026.02.01.26345312

**Authors:** Rucha SH. Wagh, Rupali U. Bawdekar, Wareed Alenaini, Caroline HD. Fall, E Louise Thomas, Jimmy D Bell, Rashmi B. Prasad, Satyajeet P. Khare, Chittaranjan S Yajnik

## Abstract

**Background:** The thin-fat Indian phenotype defined as lower lean mass and higher adiposity compared to Europeans, is present at birth and progresses throughout the lifecourse. While this increases cardiometabolic risk from a young age, there is scant information on the individual contributions of subcutaneous and visceral abdominal fat to cardiometabolic risk in young Indians. Western population-based studies link visceral adiposity (VAT) rather than abdominal subcutaneous adiposity (ASAT) to cardiometabolic disease. However, some South Asian Indian population-based studies have reported an association of ASAT with cardiometabolic risk. Whether this association persists in young South Asian populations remains unclear. We investigated associations of ASAT and VAT with cardiometabolic risk markers in young rural Indian adults.

**Methods:** We quantified ASAT and VAT using MRI at 18-years of age in 310 men and 280 women from the Pune Maternal Nutrition Study birth cohort. Sex-specific multiple regression models examined cross-sectional (18-years) and prospective (24-years) associations of ASAT and VAT with a range of cardiometabolic risk markers, and also with change in risk (delta) between two time points; adjusting for obesity indices and birth size.

**Findings:** In regression models, ASAT was strongly associated with more adverse cardiometabolic profiles (higher glucose, lipids, blood pressure, leptin, CRP, leukocyte count, and lower adiponectin and Matsuda Index), consistently showing higher effect estimates than VAT in both sexes (beta ranging from 0.45 to 0.88 in ASAT vs 0.13 to 0.18 in VAT, p<0.05). ASAT at 18-years remained significantly associated with levels of cardiometabolic risk factors at 24-years, and also with the change in their level from 18-years. These associations were independent of obesity and birth size.

**Interpretation:** Our findings suggest a possible role for ASAT in increasing cardiometabolic risk in young Indians, raising a possibility of novel adipocyte pathobiology. Despite an observational design, consistency of associations across time (18-and 24-years) and temporality are compatible with a causal hypothesis.

**Funding:** CSY: Wellcome Trust, UK; the MRC, UK; and the DBT, India. RP: VINNOVA and VR. RSHW: SRF from the CSIR, India

**Research highlights:** *Evidence before this study:* Intrauterine undernutrition programs a distinct thin-fat body composition in South Asians: low birth weight and lean mass with preserved adiposity. This phenotype is associated with elevated susceptibility to cardiometabolic risk at young age and lower BMI compared to Europeans. Western adult data asserts that central adiposity-related cardiometabolic risks are driven by visceral adipose tissue (VAT) rather than by abdominal subcutaneous tissue (ASAT). Previous studies, including ours, have raised the possibility that ASAT is also associated with increased cardiometabolic risk in adult South Asian Indians. There are few imaging studies in young Indians.

*Added value of this study:* In a well-characterised rural pre-conceptional birth cohort followed into young adulthood, we show that ASAT measured by MRI at 18 years is more strongly associated with cardiometabolic risk factors at both 18 and 24 years than VAT, which showed only weak associations. These findings suggest that ASAT may mediate the unexpectedly high metabolic burden observed in these thin, young rural Indian adults.

*Implications of all the available evidence:* Clinicians and policymakers should not overlook abdominal subcutaneous fat in risk stratification of young and lean Indians. Further research is warranted to clarify the developmental origins and cellular-molecular basis of this observation

Graphical Abstract
Population-specific pattern of cardiometabolic risk and abdominal fat depots. Western populations stress visceral adipose tissue (VAT) as the primary pathogenic abdominal fat depot. In contrast, our MRI-based analyses in young, lean rural Indians demonstrate stronger and broader associations of abdominal subcutaneous adipose tissue (ASAT) with cardiometabolic risk markers. This conceptual schematic highlights a context-dependent abdominal fat risk hierarchy, potentially shaped by genetics, early-life programming and population-specific adipose tissue biology. Red color denotes risk associations while green denotes protective effect.

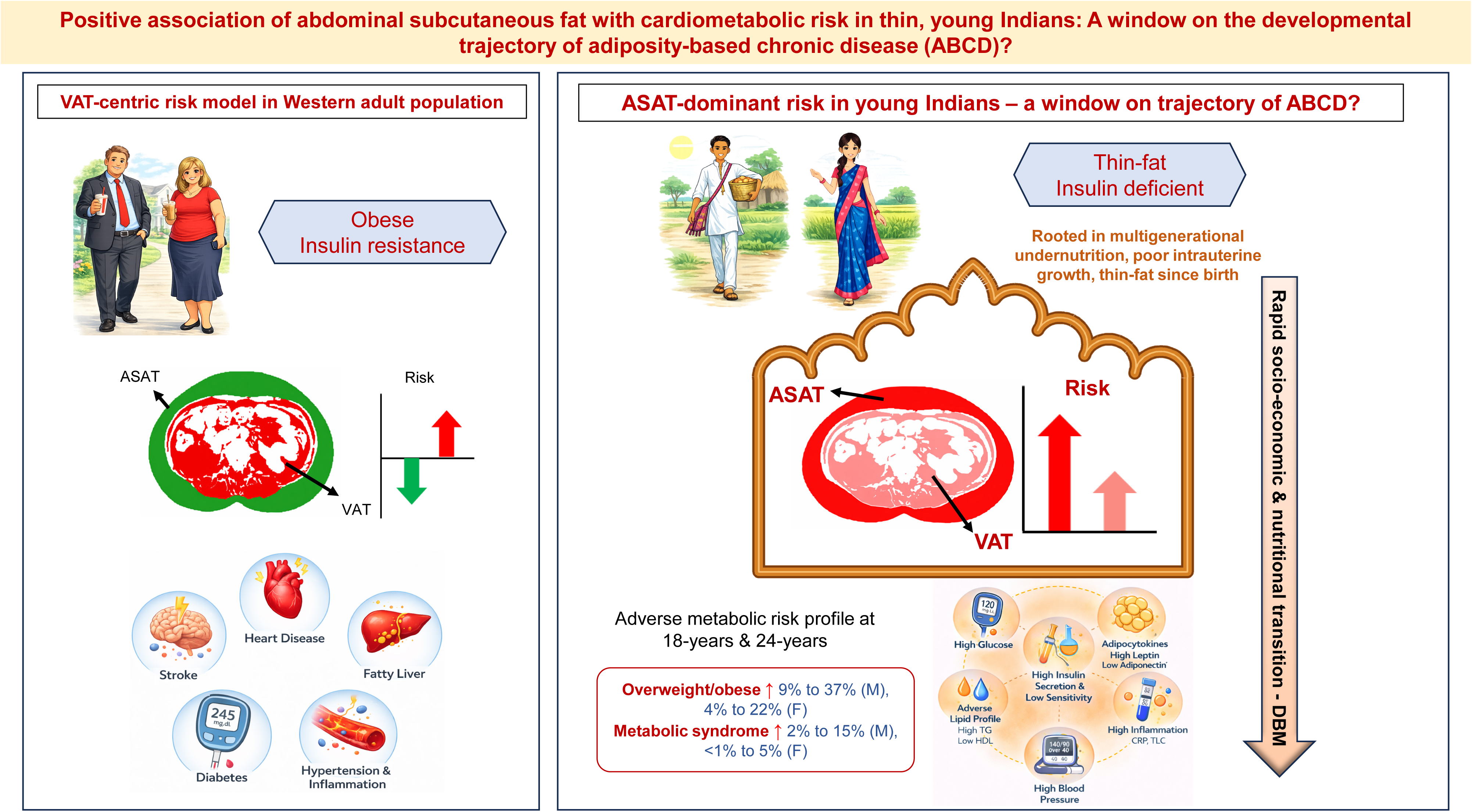

## INTRODUCTION

Obesity is increasing rapidly in low- and middle-income countries such as India, the latter with a prevalence of ∼23% [1, 2], attributed to a rapid nutritional transition from multigenerational undernutrition to overnutrition, resulting in a double burden of malnutrition[1, 2]. Body mass index (BMI), the conventional obesity metric, is a crude proxy for adiposity and provides little information on body composition [3] or adipose tissue-related organ dysfunction. This has recently been addressed by the 2025 Lancet Diabetes & Endocrinology Commission on clinical obesity, which recommended that obesity be confirmed by direct- or anthropometric measures of body composition in addition to BMI[4]. This reflects a broader conceptual shift toward viewing obesity as a disorder of adipose tissue mass, distribution, and function, reflected in the new terminology of Adiposity-Based Chronic Disease (ABCD) -rather than a disorder defined by excess weight alone[5, 6].

From the DOHaD perspective, this shift is especially relevant to South Asian Indians, whose ‘thin-fat’ phenotype from birth (thinner but relatively adipose compared to white Europeans) explains their heightened cardiovascular risk at a younger age and lower BMI. This ‘thrifty phenotype’ is ascribed to developmental programming during a nutritionally constrained intrauterine environment [7], in addition to genetic susceptibility [8]. This phenotype has, to date, been characterised largely through anthropometry and whole-body DXA[9, 10]. MRI-based deep phenotyping, resolving abdominal fat into visceral (VAT) and subcutaneous (ASAT) compartments, allows extension of the thin-fat phenotype to depot-specific risk factors for cardiometabolic disease.

In Western populations, VAT is consistently associated with insulin resistance, dyslipidaemia, type 2 diabetes, and inflammation[11], whereas ASAT is not associated with cardiometabolic risk[12, 13]. However, in Asian Indians, ASAT and VAT have both been linked to adverse cardiometabolic risk, especially in younger adults[14–16], whereas in older adults, the association appears to be restricted to VAT[17, 18]. This heterogeneity may reflect differences in age and metabolic status across study populations, in addition to the differences in the imaging and anthropometric methods[14–18]. We had an opportunity to examine associations between MRI-measured ASAT and VAT at 18 years of age and cardiometabolic risk factors in the Pune Maternal Nutrition Study (PMNS) rural birth cohort, at 18 and 24 years of age in apparently healthy, lean, young Indian adults[19]. We hypothesised that both ASAT and VAT would be associated with cardiometabolic risk markers in these young and thin adults.

## METHODS

### Overview of the cohort

The PMNS is a preconceptional birth cohort established in 1993 in six villages near Pune, India[19]. Of 2466 married non-pregnant women (F0 generation) invited to participate, 797 who became pregnant and had a singleton pregnancy confirmed by ultrasound (<21 weeks’ gestation) were enrolled with their spouses and offspring (F1 generation). Parents and offspring have since been followed six-yearly, for detailed measurements of body size, body composition and cardiometabolic risk factors. This analysis uses the two most recent F1 follow-ups: 18 years (2012-2014) and 24 years (2019-2025, mean 25.2 years). The 24 years follow-up was prolonged by the COVID-19 pandemic, resulting in a wider age range; the label 24 years denotes visit sequence rather than precise age.

At 18 years, 663 participants were examined, of whom 599 underwent abdominal MRI. Relevant data was available on 590 (310 men, 280 women) (Supplementary Figure S1). Of these, 551 (290 men, 261 women) attended the 24 years follow-up. All participants arrived in the Diabetes Research Centre on the evening before the day of measurements. They were provided with a standard dinner, and fasted overnight. MRI and other measurements were done on the next day.

### Anthropometric measurements

Height, weight, and waist and hip circumferences were measured by trained staff using standardised methods (Supplementary Table S1). BMI was calculated as weight(kg)/height(m)².

### MRI measurements at 18 years

Magnetic Resonance Imaging was performed using a 1.5T Siemens MAGNETOM Harmony scanner (Siemens Corporation, Munich, Germany). Axial abdominal images were acquired using a T1-weighted spin-echo sequence (repetition time 100 ms, echo time 7.49 ms, one-half excitation), with 10-mm slice thickness and an 11-mm inter-slice gap. For each participant, three slices were obtained, centred at the L4–L5 intervertebral disc, with a total acquisition time of 36 seconds. The images were quantified in collaboration with Research Centre for Optimal Health, School of Life Sciences, Westminster University, London, UK. ASAT and VAT were quantified using Slice-O-Matic (Tomovision, Montreal, QC, Canada). Segmentation followed an established protocol, which combines SliceOMatic’s image-morphology tools to delineate adipose tissue boundaries with manual expert editing to assign each region as ASAT or VAT. This workflow has demonstrated high reproducibility in previous studies, with coefficients of variation typically <1.1% for ASAT and 1.6% for VAT [20]. The same trained operators and identical segmentation workflow were used in the present study, ensuring consistency with these previously validated methods.

### Biochemical measurements

At 18- and 24-years, fasting and post-load (30- and 120-minutes) glucose and insulin concentrations were measured during a 75g oral glucose tolerance test (OGTT). Insulin sensitivity was estimated using the Matsuda Index[21], and early insulin secretion using the insulinogenic index[22]. Disposition index, a measure of insulin secretion adjusted for prevailing insulin sensitivity, was calculated as the product of the insulinogenic and the Matsuda indices. Fasting plasma triglycerides, total and HDL-cholesterol concentrations were measured using standardised enzymatic assays (Hitachi 902, Roche Diagnostics, GmbH, Germany), LDL-cholesterol was derived using the Friedewald equation, and the triglyceride:HDL-cholesterol ratio was calculated as a surrogate marker of metabolic dysfunction. Inflammatory markers including high sensitivity C-reactive protein and total leukocyte count were measured by high-sensitivity ELISA (hsCRP, United Biotech. Mountain View, CA, USA) and Beckman Coulter Analyzer (AC.T diff Analyzer, Miami, Florida, USA) respectively. Circulating leptin and adiponectin were measured only at 18-years by ELISA (Alpco Diagnostics, Salem, NH 03079).

## Definitions

The following definitions were applied at both 18- and 24-years. BMI categories followed WHO criteria (underweight <18.5; normal 18.5-25; overweight/obese >25 kg/m²)[23]. Central obesity: waist circumference ≥90 cm (men), ≥80 cm (women). Prediabetes (ADA 2014)[24]: impaired fasting glucose (IFG) and/or impaired glucose tolerance (IGT) (fasting glucose ≥5.6 mmol/L and/or 120-min glucose 7.8–11.0 mmol/L). Dyslipidaemia: triglycerides >1.69 mmol/L and/or HDL-cholesterol <1.03 (men)/<1.29 (women) mmol/L. Hypertension: BP ≥130/85 mmHg. Metabolic syndrome (IDF criteria)[25]: central obesity plus 2 or more of the following: triglycerides >1.69 mmol/L, HDL-cholesterol <1.03/<1.29 mmol/L, BP ≥130/85 mmHg, fasting glucose ≥5.6 mmol/L.

Socioeconomic status (SES) was assessed using the Standard of Living Index system of the National Family Health Survey-4[26]. History of smoking and alcohol consumption was recorded.

## Statistical analysis

Data are presented as median (25th–75th percentile), skewed variables were log-transformed. Within each sex (n = 310 males, n = 280 females), all exposure and outcome variables were converted to standardized residuals (mean 0, SD 1) using regression after adjusting for age and socioeconomic status (SES). Age and SES were selected as the primary covariates to construct a parsimonious model that adequately controls for key demographic confounding while preserving statistical power, given our sample size. The standardized residuals, reflecting variance independent of age and SES, were then used in subsequent regression models. Linearity of both VAT and ASAT with outcomes was confirmed via LOWESS plots (Supplementary Figure S2).

Associations between 18-year VAT and ASAT, and cardiometabolic markers at 18 and 24 years were examined for each sex using multivariable linear regression. VAT and ASAT were moderately correlated (Spearman’s ρ = 0.43 overall; 0.49 men, 0.37 women, p<0.05 for all), both were entered into the same model to estimate the independent effect of each depot (acceptable collinearity as shown by VIF ranging from 1.2-1.4; and Supplementary Table S2). The general model is presented as,

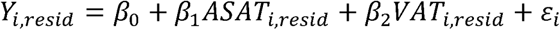

where,

β□ and β□ give the SD change in the outcome per 1 SD change in ASAT or VAT, respectively, with the other depot held constant. A Wald test compared the ASAT versus VAT coefficients for each outcome.

To examine whether 18-year ASAT and VAT predicted subsequent change in cardiometabolic risk, independent of their cross-sectional/prospective associations, we tested their associations with change in outcomes between 18- and 24-years (Δresid = resid□□ Y − resid □□Y) using linear regression.

In addition, we investigated the effect of adjustments for obesity (BMI, waist circumference, and height); and early life growth (birthweight, birth length, and head circumference). To further test robustness of our findings, we stratified the participants into high and low VAT:ASAT ratio groups (median split), and compared cardiometabolic outcomes using a Wilcoxon test within each sex.

*Categorical outcomes* Metabolic syndrome was modelled using Firth’s penalised logistic regression to account for small-sample correction (12/590) at 18-years, and logistic regression at 24-years. Prediabetes was analysed using logistic regression at both visits.

A nominal p<0.05 was considered significant; multiple-testing correction (Benjamini-Hochberg FDR) was applied per exposure within each sex-specific analysis (16 outcomes at 18 years, 12 at 24 years).

All the analyses were performed using SPSS v26 and R v4.4.1.

## Ethics approval and consent to participate

The study was approved by local village leaders and the Ethics Committee of the KEM Hospital Research Centre (KEMHRC/VSP/Dir. Off/EC12465). An informed consent form was signed by all participants. Ethical approvals pertaining to this secondary analysis were obtained from the KEM Hospital ethics committee: KEMHRC/RVM/EC/1037 and SIU: EC/NEW/IND/2024/MH/0477.

## RESULTS

### Characteristics of participants (Table 1)

Of the 663 participants in the 18-year follow-up, 590 (310 men and 280 women) had complete abdominal MRI data and were analysed (Supplementary Figure S1). They were broadly similar to those without MRI data, except that included men were slightly older, while included women were marginally younger and had higher 120-min plasma glucose levels (Supplementary Table S3).

**Table 1:** Characteristics of the participants at 18-years.

|  | **18-year** |  |  |  | **24-year** |  |  |  |
|---|---|---|---|---|---|---|---|---|
| **Measures** | **Men (n=310)** |  | **Women (n=280)** |  | **Men (n=290)** |  | **Women (n=261)** |  |
| Age (years) | 18.2 | (17.9, 18.6) | 17.6 | (17.2, 18.1) | 26.6 | (26.0-27.2) | 23.8 | (23.3-27.7) |
| Height (cm) | 169.6 | (165.5, 173.7) | 157.0 | (153.3, 160.7) | 170.6 | (167.2-175.0) | 156.8 | (152.8-160.2) |
| Weight (kg) | 55.49 | (49.39, 62.02) | 44.8 | (41.3, 49.9) | 69.6 | (59.3-77.8) | 51.3 | (44.4-59.7) |
| BMI | 19.0 | (17.2, 21.3) | 18.7 | (16.8, 20.2) | 26.6 | (26.0-27.2) | 23.8 | (23.3-27.7) |
| BMI categories<br>Underweight (<18.5 kg/m<sup>2</sup>)<br>Overweight/Obese (>25 kg/m<sup>2</sup>) | 127<br>28 | (41.1%)<br>(9.1%) | 153<br>12 | (54.4%)<br>(4.3%) | 39<br>108 | (13.4%)<br>(37.2%) | 81<br>58 | (31.1%)<br>(22.4%) |
| Waist circumference (cm) | 71.0 | (66.5, 77.5) | 67.5 | (63.3, 71.8) | 86.9 | (77.3-93.9) | 74.4 | (67.9-82.0) |
| Central Obesity<br>(men>90cm, women>80cm) | 19 | (6.1%) | 14 | (5%) | 111 | (39%) | 78 | (31%) |
| MRI ASAT (cm²) | 217.6 | (152.2, 363.8) | 311.2 | (219.3, 442.6) | - | - | - | - |
| MRI VAT (cm²) | 194.5 | (158.5, 242.3) | 164.7 | (132.0, 204.3) | - | - | - | - |
| MRI total abdominal area (cm²) | 408.5 | (325.6, 608.8) | 473.8 | (365.9, 623.9) | - | - | - | - |
| MRI ASAT/VAT ratio | 1.2 | (0.9, 1.8) | 1.9 | (1.4, 2.7) | - | - | - | - |
| Fasting glucose (mmol/l) | 5.39 | (5.17, 5.61) | 5.17 | (4.94, 5.33) | 5.22 | (4.94-5.50) | 5.00 | (4.72-5.27) |
| 120-min glucose (mmol/l) | 6.11 | (5.33, 7.01) | 6.33 | (5.50, 7.11) | 6.27 | (5.27-7.10) | 5.88 | (5.16-6.72) |
| Prediabetes (IFG+IGT) | 124 | (40%) | 54 | (19.3%) | 87 | (30.6%) | 44 | (17.1%) |
| Diabetes | 0 | 0 | 0 | 0 | 3 | (1%) | 1 | (<1%) |
| Insulinogenic index | 1.70 | (1.39, 2.07) | 1.69 | (1.36, 2.07) | 2.02 | (1.58-2.38) | 1.95 | (1.65-2.41) |
| Matsuda index | 13.2 | (9.46, 19.12) | 11.76 | (8.81, 15.74) | 3.60 | (2.57-6.44) | 4.61 | (3.32-6.56) |
| Disposition Index | 4.31 | (3.81, 4.83) | 4.12 | (3.71, 4.57) | 7.33 | (4.24-12.87) | 9.02 | (5.58-14.34) |
| Systolic BP (mm Hg) | 112 | (106, 119) | 106 | (100, 110) | 114 | (107-121) | 104 | (98-111) |
| Diastolic BP (mm Hg) | 58 | (53, 64) | 61 | (57, 65) | 69 | (62-75) | 66 | (61-71) |
| Triglycerides (mmol/l) | 0.69 | (0.53, 0.86) | 0.60 | (0.49, 0.75) | 0.97 | (0.72-1.41) | 0.62 | (0.51-0.86) |
| Total cholesterol (mmol/l) | 3.14 | (2.79, 3.59) | 3.44 | (3.03, 3.79) | 4.21 | (3.78-4.81) | 3.90 | (3.44-4.29) |
| HDL-cholesterol (mmol/l) | 0.98 | (0.85, 1.09) | 1.09 | (0.96, 1.24) | 0.98 | (0.85-1.14) | 1.19 | (1.04-1.32) |
| LDL-cholesterol (mmol/l) | 1.84 | (1.54, 2.21) | 2.00 | (1.67, 2.36) | 2.69 | (2.33-3.19) | 2.33 | (1.99-2.77) |
| Metabolic syndrome | 11 | (3.5%) | 1 | (<1%) | 44 | (15%) | 13 | (5%) |
| Leptin (ng/mL) | 2.7 | (1.2, 6.7) | 15.8 | (10.1, 25.1) | - | - | - | - |
| Adiponectin (ng/mL) | 4.7 | (3.7, 5.9) | 5.1 | (4.0, 6.5) | - | - | - | - |
| Total leucocyte count (10<sup>3</sup>/µL) | 5.8 | (5.1, 6.6) | 6.3 | (5.6, 7.4) | 6.5 | (5.6-7.7) | 6.7 | (5.7-8.0) |
| C-reactive protein (mg/L) | 0.26 | (0.11-0.84) | 0.21 | (0.08-0.51) | 0.90 | (0.36-2.43) | 0.86 | (0.28-2.33) |

Data is presented as median (25<sup>th</sup>-75<sup>th</sup>) or N (%) where appropriate.

At the 18-year follow-up, the participants were overall lean (median BMI 19.0 kg/m^2^ in men, 18.1 kg/m^2^ in women). More than 40 % of men and half of the women were underweight, while overweight/obesity was uncommon (9.1% men and 4.3% in women, Table 1). Central obesity, defined by higher WC, was present in 6.1% of men and 5% of women. Fewer than 1% reported a history of smoking and alcohol consumption. Even so, prediabetes was prevalent in 40% of men and 19.2% of women, driven by IFG in men (28.4 % vs 8.5%) and IGT in women (8.2% vs 4.2%). Dyslipidaemia affected about a third of participants (32% in men and 38.4 % of women). No participant was hypertensive, and 12 (2%) met the IDF criteria for metabolic syndrome. MRI revealed clear sex differences in fat distribution. Women, compared to men, had a greater total abdominal fat (473.8 vs 408.5 cm^2^), ASAT (311.2 cm^2^ vs 217.6 cm^2^), and a higher ASAT-to-VAT ratio (1.9 vs 1.2), while men had a higher VAT (194.5 cm^2^ vs 164.7 cm^2^) (Table 1).

By the 24-year follow-up (551 participants with complete data; 290 men and 261 women), the metabolic profile had worsened markedly (Table 1). While fewer than before were still underweight (13% of men and 31% of women), overweight/obesity rose to 37% in men and 22% in women and central obesity to 39% in men and 31% in women. Smoking and alcohol consumption also increased to 5% and 10% respectively, mostly in the men. Dyslipidaemias roughly doubled (62% in men and 68% in women). Hypertension appeared, mainly in men (11% vs 2% in women), and metabolic syndrome rose to 15.2% in men and 5.1% in women. Glucose intolerance (including diabetes) was present in 31% of men and 17% of women.

### ASAT has stronger associations with cardiometabolic risk markers than VAT

Given established sex differences in body composition, all analyses were stratified by sex. As VAT and ASAT were moderately correlated, their age-and SES-adjusted standardized residuals were entered simultaneously in each model, so that the estimates reflect the independent, mutually adjusted association of each depot.

#### Cross-sectional associations (18 years)

In men, VAT was positively associated with systolic blood pressure, and negatively with the Matsuda index. ASAT was positively associated with 120-min glucose, triglycerides, total and LDL-cholesterol concentrations, TG:HDL-c ratio, systolic and diastolic blood pressure, leptin, hsCRP, and total leukocyte count, and negatively associated with the Matsuda index and circulating adiponectin (Figure 1, Supplementary Table S4). In women, VAT was negatively associated with HDL-c concentrations. ASAT showed positive associations with systolic and diastolic blood pressure, triglycerides, total and LDL-cholesterol concentrations, TG:HDL-c ratio, leptin, hsCRP, and leukocyte count, and negative with the Matsuda index and adiponectin (Figure 2, Supplementary Table S4). In both sexes, associations were stronger for ASAT than for VAT, as supported by Wald tests (e.g., Matsuda Index in men ASAT vs VAT, Z = -4.39, p<0.001).

**Figure 1.**
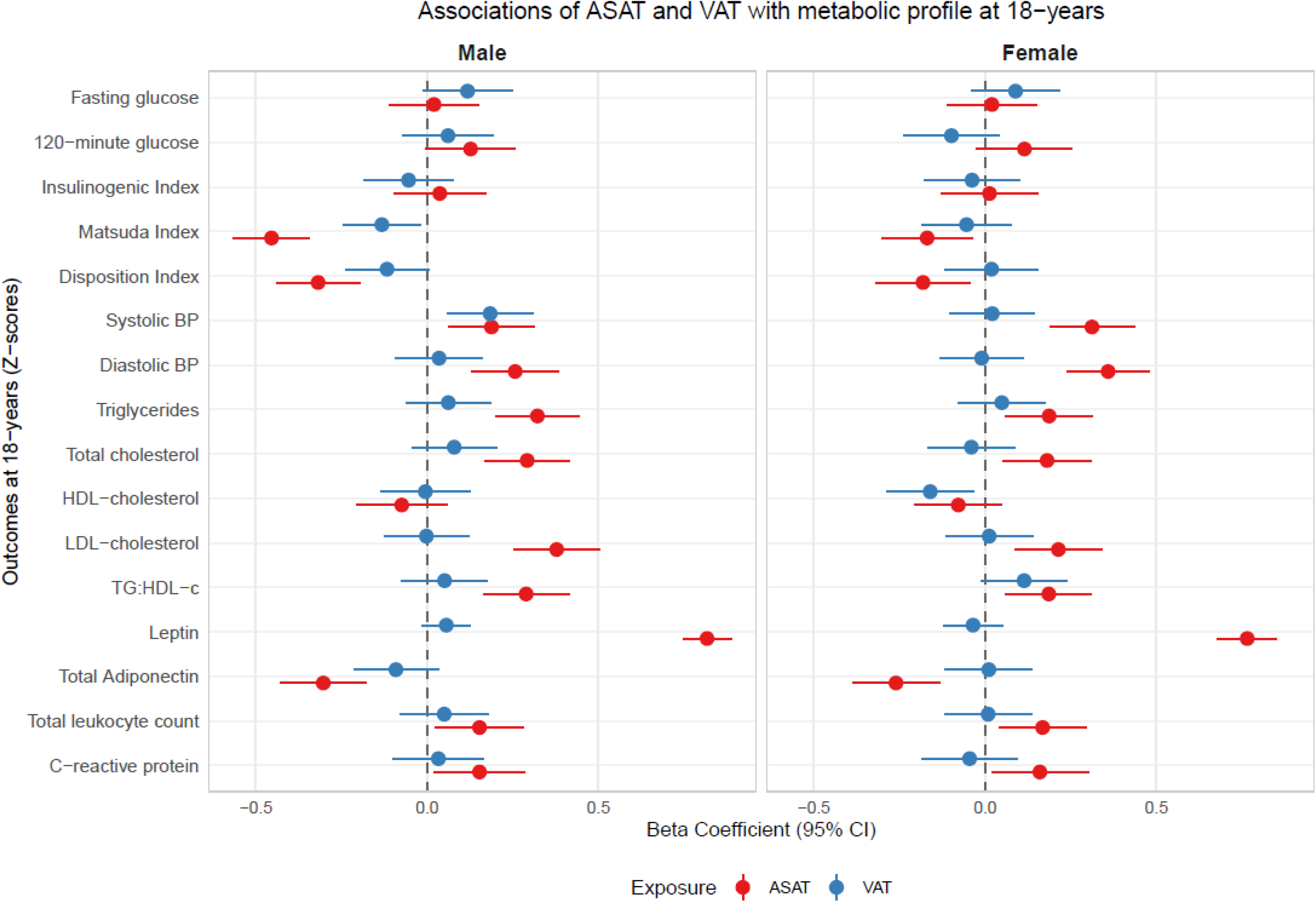
Associations between VAT, ASAT, and the cardiometabolic profile of the participants at 18-years adjusted for age and SES, and separately for sexes. Line diagram showing the associations (beta estimates) between VAT, ASAT, and the metabolic profile of the participants separately for sexes – (a) Men, (b) Women.

**Figure 2.**
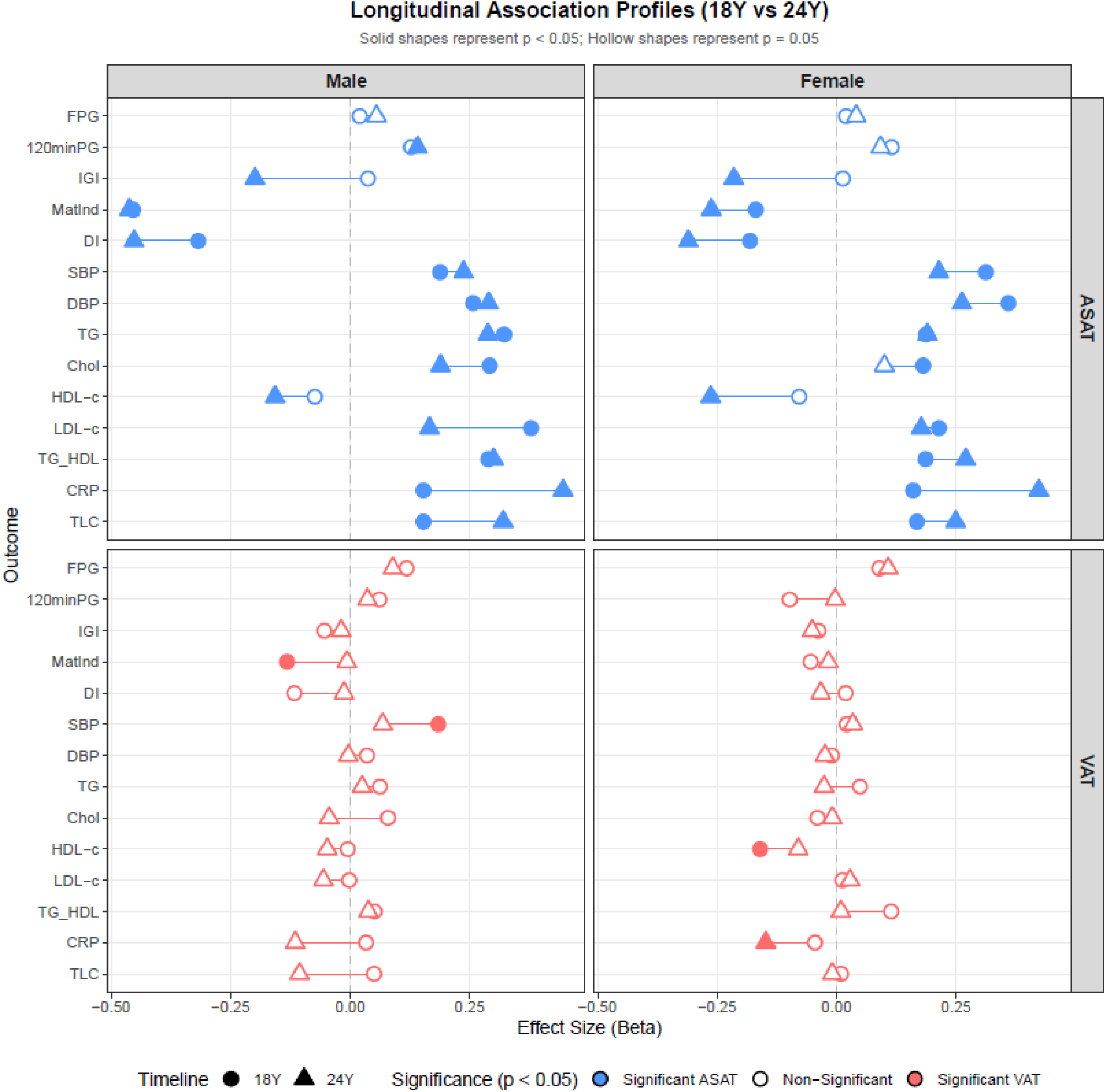
Associations between VAT and ASAT at 18-years, and the cardiometabolic profile of the participants at 18- and 24-years adjusted for age and separately for sexes. Line diagram showing the associations (beta estimates) between VAT and ASAT at 18-years, and the metabolic profile of the participants at 18-years (denoted by a circle) and ‘24-years’ (denoted by triangle) for markers available at both points separately for sexes – (a) Men, (b) Women. Significant associations are indicated as solid shapes and non-significant as hollow shapes.

#### Prospective associations (18 to 24 years)

ASAT at 18-years remained consistently and significantly associated with cardiometabolic risk markers at 24-years in both sexes, while associations for VAT were attenuated (Figure 2, Supplementary Table S5). We also assessed the associations of ASAT and VAT at 18-years with the change in the cardiometabolic risk markers between 18-and 24-years (Figure 3, Supplementary Table 6): In men, higher ASAT was significantly associated with the increase in CRP and total leukocyte concentrations, and the decrease in insulinogenic and disposition indices, and LDL-c concentrations, while higher VAT with increase in Matsuda index, and decrease in total leukocyte concentrations. In women, higher ASAT was significantly associated with increase in CRP and decrease in HDL-c concentrations, while VAT showed no significant associations. These findings indicate that depot-specific adiposity predicts the subsequent trajectory of cardiometabolic markers.

**Figure 3.**
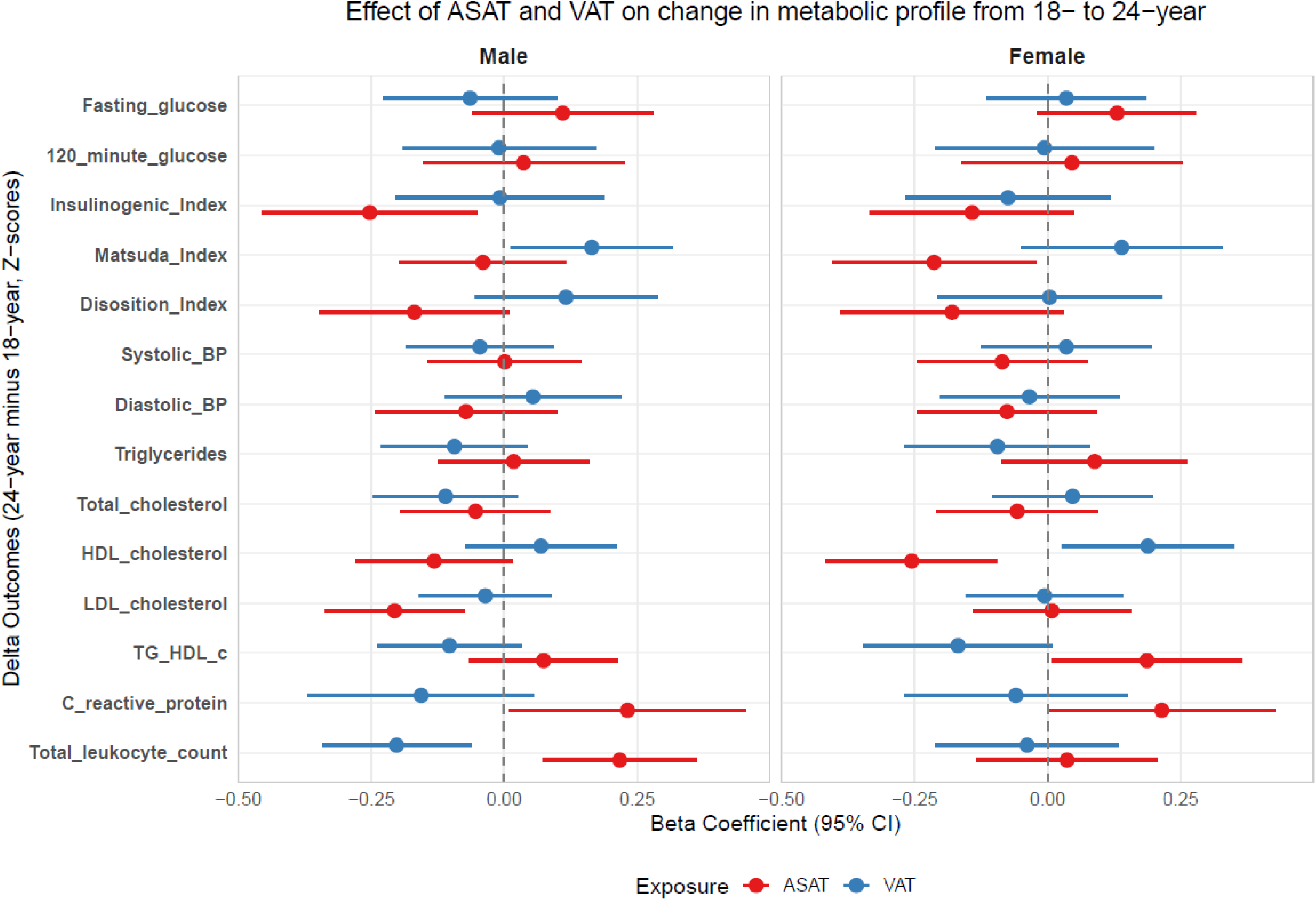
Associations between VAT and ASAT at 18-years, and change in the cardiometabolic profile of the participants between 18- and 24-years adjusted for age separately for sexes. Line diagram showing the associations (beta estimates) between VAT and ASAT at 18-years, and the change in the cardiometabolic profile (delta) of the participants between 18- and ‘24-years’ for markers available at both points separately for sexes – (a) Men, (b) Women.

Additional adjustment for height (Supplementary Table S7) or birth measures (Supplementary Table S10-S12) did not alter these findings, but BMI and WC moderately attenuated the associations both cross-sectionally and prospectively (Supplementary Tables S8, S9). Metabolic syndrome related variables were higher in the low VAT:ASAT ratio group, indicating a predominant role for ASAT (Supplementary Table S13).

#### Categorical outcomes

ASAT, but not VAT, was associated with metabolic syndrome both at 18- and 24-year (Supplementary Figure S3). Neither depot was associated with prediabetes at either of the timepoints (Supplementary Figure S3).

## DISCUSSION

The PMNS is a population-based study of a sizeable number of apparently healthy, lean, young rural Indian adults belonging to a well characterised birth cohort set up to study the DOHaD paradigm. We used MRI to quantify abdominal fat compartments and studied their associations with cardiometabolic risk markers. Traditionally, such a population would be expected to have a low burden of cardiometabolic risk. However, more than a quarter of these young adults already had glucose intolerance at 18-years, which we have previously linked to poor intrauterine growth and higher maternal pregnancy glycaemia (within normal range) [27]. This presents an early signal that the metabolic trajectory in this cohort was shaped well before adulthood. At 18-years of age (when the MRI was performed), a sizeable proportion were classified as underweight, however from 18- to 24-years of age, overweight/obesity increased in both men and women, highlighting the evolving double burden of malnutrition (Table 1). We found significant associations of ASAT with insulin resistance indices, higher blood pressure, abnormal lipid levels, adipokines, and inflammatory markers in both sexes. On the other hand, VAT was associated with fewer risk factors. ASAT at 18-years was significantly associated with cardiometabolic profile at 24-years, and was similarly associated with metabolic syndrome at both timepoints, while VAT did not. These associations persisted independent of overall body size at age 18 and birth measures. Low-VAT:ASAT ratio group showed a more adverse metabolic profile confirming a predominant role for ASAT. Higher ASAT at 18-years also predicted a subsequent worsening of several cardiometabolic markers over the next six years: increased inflammation in men and women and, declining insulin secretory capacity in men, whereas VAT showed few such associations. Rather than interpreting these findings simply as a chance departure from the VAT-centric model in Western adult populations[11], we would like to suggest that in young, thin, rural Indian adults, ASAT is associated with a more adverse cardiometabolic profile, and that the adverse influence continues over the duration of our follow-up. A longer influence will be evident only after further follow-up.

These results expand the concept of the detrimental thin-fat phenotype originally based on whole-body fat percent to MRI-measured abdominal subcutaneous and visceral fat depots. Given the considerable literature in Europeans highlighting a strong role for visceral fat in increasing cardiometabolic risk, our findings of prominent role for subcutaneous fat are somewhat unexpected. However, a sizeable literature in South Asian adults and in patients with diabetes has reported an association of subcutaneous fat with dyslipidaemia, hypertension, and diabetes [14–18]. It should also be noted that the neonatal thin-fat phenotype defined by skinfolds and by MRI imaging is predominantly driven by the subcutaneous fat [3, 7, 28], and continues into childhood [29, 30]. It is interesting that the thin-fat Indian neonate also had higher leptin and lower adiponectin concentrations in the cord blood compared to the much heavier English babies highlighting a dysfunction of subcutaneous adipose tissue. Histological studies of adipocytes show larger, hypertrophic cells and smaller number, especially, of small adipocytes in abdominal subcutaneous adipose tissue of South Asians compared with Europeans[31–34]. Transcriptomic analysis in subcutaneous adipocytes of young South Asians in UK showed reduced expression of genes involved in lipogenesis (triglyceride uptake, turnover, and storage) and insulin signalling compared to white Europeans. Multi-omics analyses in Indians in India show overlapping pro-inflammatory and lipogenic gene expression signatures in both ASAT and VAT[31, 35]. Overall, these studies suggest a characteristic adipose tissue pathobiology in Indians compared to white Europeans.

The adipose tissue expandability hypothesis proposes that each individual has a finite capacity to expand subcutaneous adipose tissue safely, and once this is exceeded, excess lipid is diverted to ectopic and visceral depots, triggering the lipotoxic cascade that underlies insulin resistance and downstream cardiometabolic disease [36]. Sniderman et al. extended this concept to South Asians, proposing that an inherently smaller subcutaneous adipose tissue compartment exhausts storage capacity at a lower threshold of overall adiposity than in White populations, resulting in an ‘overflow’ of lipid into visceral and other ectopic sites [37]. Our finding that ASAT, more than VAT, had the stronger and more consistent association with cardiometabolic risk factors in young adults adds a developmental dimension: if subcutaneous adipose tissue expandability in South Asians is itself constrained from early life, as supported by higher leptin and lower adiponectin concentrations in the cord blood of Indian neonates compared to white Europeans, then the ASAT depot may function less as a protective buffer and more as a marker of adiposopathy. This reframes the debate around which fat depot is ‘worse’ and instead positions ASAT as a developmentally sensitive readout of an individual’s underlying capacity for healthy fat storage, a capacity plausibly shaped in utero and detectable well before the metabolic consequences become clinically overt. We found both ASAT and VAT were associated with cardiometabolic risk in men, whereas ASAT was the dominant correlate in women. These differences likely reflect the influence of sex hormones on adipose tissue distribution and metabolism during puberty[38].

Strengths of this study include an MRI-based quantification of abdominal adipose depots in 590 participants from a well-characterised birth cohort, comprehensive cardiometabolic phenotyping, and assessment at a young age before overt disease and treatment effects, thereby reducing reverse causality. This study adds a depot-specific resolution to the thin-fat concept in young Indians. Limitations include an observational design which precluded causal assertions, though the consistent associations with a range of cardiometabolic risk markers both cross-sectionally and prospectively (at 24-years) raise the possibility of causal associations. We have not shown dietary intake and physical activity data in this paper for simplicity. Other limitations include the lack of data on superficial and deep ASAT compartments as well as gluteofemoral depots, and use of epidemiological rather than gold-standard measures of insulin action (insulin clamps).

In young, lean individuals from rural India, abdominal subcutaneous rather than visceral adiposity is linked with adverse cardiometabolic profile. Our findings are best interpreted in a lifecourse model starting from birth and continuing into young adult age indicating that a long-term metabolic blueprint is established early in life, well before overt weight gain or clinical obesity emerge. Taken together, these findings argue for age- and ethnicity-specific approaches to cardiometabolic risk assessment, prevention, and intervention in South Asian populations. Further mechanistic studies that characterise superficial from deep ASAT and VAT, are needed to clarify the developmental, cellular, and molecular pathways underlying this depot-specific programming and its long-term metabolic consequences.

## Supporting information

All the supplementary tables and figures

## Declarations

### Data Availability and Code Availability statements

The dataset supporting the conclusions of this article is available for additional analyses by applying to the corresponding author with a 200-word plan of analysis. Data sharing is subject to KEM Hospital Research Centre administrative approvals and permission from the Government of India’s Health Ministry Screening Committee. The analysis code has been uploaded in the github repository and is accessible using the URL (https://github.com/RuWa3/PMNSmain_MRI-Feb-2026/tree/main).

### Funding

The PMNS was funded by the Wellcome Trust, UK (038128/Z/93, 059609/Z/99, 079877/Z/06/Z, 098575/B/12/Z and 083460/Z/07/Z); the Medical Research Council, UK (MR/J000094/1); and the Department of Biotechnology, Government of India (BT/PR-6870/PID/20/268/2005). Rashmi Prasad has received funding support from VINNOVA (2023-04234 ) and VR (2021-02623). RW is supported by a senior research fellowship from the Council of Scientific and Industrial Research, India

## Acknowledgements

We thank the participants, collaborators, funding agencies and the Diabetes Unit staff for their contribution towards this study.

## Author Contributions Statement

CSY and CHDF conceptualised the study. RSHW was involved in statistical analysis. RB and WA contributed to MRI image data cleaning and image segmentation. ELT and JDB set up and supervised the MRI image segmentation. CSY and RSHW prepared the manuscript. SK and RP were involved in the PhD supervision (RSHW) and critical review of the manuscript. All authors were involved in the review of the manuscript, and approved the final version of the manuscript. CSY is the guarantor of this work and, as such, had full access to all the data in the study and takes responsibility for the integrity of the data and the accuracy of the data analysis

## Declaration of interests

The authors declare that they have no conflict of interests.

### Prior Presentation

A non-peer-reviewed version of this article was submitted to the medRxiv preprint server (doi: https://doi.org/10.64898/2026.02.01.26345312) on February 27, 2026

**Supplementary Figure S1. STROBE diagram of the PMNS cohort**

**Supplementary Figure S2.** LOWESS plots assessing linearity between outcome-exposure pairs. All outcomes and exposures are expressed as standardized residuals adjusted for age and socioeconomic status.

**Supplementary Figure S3**. Odds ratios for glucose intolerance at 18-years (124/310 men and 54/280 women) as well as 24-years (90/285 men and 45/257 women); and metabolic syndrome at 18-years (12/590) as well as ‘24-years’ (44/287 men and 13/256 women) as outcomes respectively. Given the small numbers of metabolic syndrome at 18-years, sex stratified analysis was not performed.

**Supplementary Figure S4. Distribution of BMI, ASAT, VAT, and ASAT/VAT ratio across South-Asian population (Native + Immigrant).** Study level means were standardised to z-scores within each phenotype before plotting across age. Cohorts – native SA (light colour): PMNS, CRISIS, CURES; migrant SA (dark colour): Immigrant SA from different UK studies (UK SA 1, UK SA 2), Mediators of Atherosclerosis in South Asians Living in America (MASALA) and Multi-Ethnic Study of Atherosclerosis (MESA), USA; The Molecular Study of Health and Risk in Ethnic Groups (MolSHARE), Canada

## Added value of this study

In a well-characterised rural pre-conceptional birth cohort followed into young adulthood, we show that ASAT measured by MRI at 18 years is more strongly associated with cardiometabolic risk factors at both 18 and 24 years than VAT, which showed only weak associations. These findings suggest that ASAT may mediate the unexpectedly high metabolic burden observed in these thin, young rural Indian adults.

## Implications of all the available evidence

Clinicians and policymakers should not overlook abdominal subcutaneous fat in risk stratification of young and lean Indians. Further research is warranted to clarify the developmental origins and cellular-molecular basis of this observation.

## Notes

### Competing Interest Statement

The authors have declared no competing interest.

### Author Declarations

The study was approved by local village leaders and the Ethics Committee of the KEM Hospital Research Centre (KEMHRC/VSP/Dir. Off/EC12465). An informed consent form was signed by all participants. Ethical approvals pertaining to this secondary analysis were obtained from KEM: KEMHRC/RVM/EC/1037 and SIU: EC/NEW/IND/2024/MH/0477

### Summary of Updates

The manuscript uses the same Pune Maternal Nutrition Study birth cohort MRI data but reframes the scientific question and adds longitudinal analyses. The original version presented the results mainly as a challenge to the visceral fat paradigm established in Western populations. In the revision, we interpret our results in the context of the thin-fat phenotype and developmental origins (DOHaD). The MRI data includes 590 18-year-old, lean, rural Indian adults born in the birth cohort. Abdominal subcutaneous adipose tissue (ASAT) showed broader and generally stronger associations than visceral adipose tissue (VAT) with cardiometabolic risk markers, and also with the risk markers at the later follow-up visit (age 24 years), whereas VAT showed fewer such associations. Additional adjustment for height and birth measures did not materially alter the findings, while adjustment for BMI and waist circumference partly attenuated them. The revision includes three main changes (1) The Discussion has been rewritten around the DOHaD and adiposity-based chronic disease frameworks, relating the findings to the thin-fat phenotype, the adipose tissue expandability hypothesis and the cohort's birth and growth characteristics. (2) We added analyses relating ASAT and VAT at age 18 to cardiometabolic markers at the later follow-up visit (age 24 years) and to change in markers between visits. (3) The additional analyses include sex-stratified, mutually adjusted models with formal Wald tests comparing ASAT and VAT estimates: sensitivity analyses adjusting for height, BMI, waist circumference and birth measures, and comparisons of high and low VAT:ASAT ratio groups.

