## Supplementary material for "Positive association of abdominal subcutaneous fat with cardiometabolic risk in thin, young Indians: A window on the developmental trajectory of adiposity-based chronic disease (ABCD)?": All the supplementary tables and figures

Index

| Sr. No. | Description | Page no. |
| --- | --- | --- |
| 1 | Supplementary table S1: Methods for anthropometry measurements in children | 2 |
| 2 | Supplementary Table S2: Variance Inflation factor (VIF) for the outcome-exposure pairs on a regression model including both VAT and ASAT as predictors | 3 |
| 4 | Supplementary Table S3: Characteristics of participants who underwent MRI and those who did not | 6 |
| 5 | Supplementary Table S4: Associations of ASAT and VAT at 18-years with cardiometabolic markers at 18-years | 7 |
| 6 | Supplementary table S5: Associations of ASAT and VAT at 18-years with cardiometabolic markers at 24-years | 8 |
| 7 | Supplementary Table S6: Associations of MRI ASAT and VAT with change in outcome (Delta) between 18- and 24-years | 9 |
| 8 | Supplementary Table S7: Associations of ASAT and VAT at 18-years with cardiometabolic markers at 18- and 24years with an additional adjustment for height at 18-years | 10-11 |
| 9 | Supplementary Table S8: Associations of ASAT and VAT at 18-years with cardiometabolic markers at 18- and 24years with an additional adjustment for BMI at 18-years | 12-13 |
| 10 | Supplementary Table S9: Associations of ASAT and VAT at 18-years with cardiometabolic markers at 18- and 24years with an additional adjustment for waist circumference at 18-years | 14-15 |
| 11 | Supplementary Table S10: Associations of ASAT and VAT at 18-years with cardiometabolic markers at 18- and 24years with an additional adjustment for birthweight | 16-17 |
| 12 | Supplementary Table S11: Associations of ASAT and VAT at 18-years with cardiometabolic markers at 18- and 24years with an additional adjustment for birth length | 18-19 |
| 13 | Supplementary Table S12: Associations of ASAT and VAT at 18-years with cardiometabolic markers at 18- and 24years with an additional adjustment for birth head circumference | 20-21 |
| 14 | Supplementary Table S13: Comparison between low and high VAT:ASAT ratio groups classified as below and above-median ratio | 22 |
| 15 | Supplementary Figure S1: STROBE flow diagram of Pune Maternal Nutrition Study | 23 |
| 16 | Supplementary Figure S2: LOWESS plots assessing linearity between outcome-exposure pairs | 24 |
| 17 | Supplementary Figure S3: Odds ratios for glucose intolerance and metabolic syndrome | 25 |

Supplementary Table S1: Methods for anthropometry measurements in children

| **Anthropometry** | | | |
| --- | --- | --- | --- |
| Measurement | Time point | Method | Least count |
| Weight | Within 72 hours of birth | Salter spring balance (Salter Abbey, Suffolk, U.K.) | Nearest 50 g |
|  | 18- and 24-years | Electronic weighing scales (ATCO Healthcare Ltd, Mumbai, India) | Nearest 10 g |
| Crown heel length (Supine) | Within 72 hours of birth and 6 monthly upto 2 years | Portable Pedobaby Babymeter (ETS J.M.B., Brussels, Belgium) | Nearest 0.1 cm |
| Standing height | 6 monthly intervals from 2- 5 years | Portable Harpenden stadiometer (Microtoise, CMS Instruments Ltd, London, UK) | Nearest 0.1 cm |
|  | 18- and 24-years | Wall-mounted stadiometer (Microtoise, CMS Instruments Ltd, London, UK) | Nearest 0.1 cm |
| Waist and Hip circumference | 18- and 24-years | Fibreglass tapes (CMS Instruments). | Nearest 0.1cm |

Supplementary Table S2: Variance Inflation factor (VIF) for the outcome-exposure pairs on a regression model including both VAT and ASAT as predictors

| outcome | sex | VIF |
| --- | --- | --- |
| Fasting glucose | Women | 1.21 |
| Fasting glucose | Men | 1.38 |
| 120-minute glucose | Women | 1.23 |
| 120-minute glucose | Men | 1.39 |
| Insulinogenic Index | Women | 1.24 |
| Insulinogenic Index | Men | 1.37 |
| Matsuda Index | Women | 1.21 |
| Matsuda Index | Men | 1.38 |
| Disposition Index | Women | 1.24 |
| Disposition Index | Men | 1.38 |
| Systolic BP | Women | 1.21 |
| Systolic BP | Men | 1.39 |
| Diastolic BP | Women | 1.21 |
| Diastolic BP | Men | 1.39 |
| Triglycerides | Women | 1.21 |
| Triglycerides | Men | 1.38 |
| Total cholesterol | Women | 1.21 |
| Total cholesterol | Men | 1.38 |
| HDL-cholesterol | Women | 1.21 |
| HDL-cholesterol | Men | 1.38 |
| LDL-cholesterol | Women | 1.21 |
| LDL-cholesterol | Men | 1.4 |
| TG:HDL-c | Women | 1.21 |
| TG:HDL-c | Men | 1.38 |
| Leptin | Women | 1.21 |
| Leptin | Men | 1.38 |
| Total Adiponectin | Women | 1.19 |
| Total Adiponectin | Men | 1.36 |
| Total leukocyte count | Women | 1.21 |
| Total leukocyte count | Men | 1.38 |
| C-reactive protein | Women | 1.24 |
| C-reactive protein | Men | 1.38 |

Supplementary Table S3: Characteristics of participants who underwent MRI and those who did not

| **Measure** | **MRI Done** | | | | **MRI Not Done** | | | |
| --- | --- | --- | --- | --- | --- | --- | --- | --- |
|  | **Men**  **(n=310)** | | **Women**  **(n=280)** | | **Men**  **(n=46)** | | **Women**  **(n=26)** | |
| **Age (years)** | **18.2** | **(17.9, 18.6)** | **17.6** | **(17.2, 18.1)** | **18.1*** | **(17.8, 18.3)** | **17.9*** | **(17.6, 18.3)** |
| Height (cm) | 169.6 | (165.5, 173.7) | 157.0 | (153.3, 160.7) | 170.6 | (166.7, 174.5) | 158.4 | (150.5, 162.2) |
| Weight (kg) | 55.5 | (49.4, 62.0) | 44.8 | (41.3, 49.9) | 55.6 | (48.8, 63.9) | 44.8 | (39.4, 47.6) |
| Body Mass Index (BMI, kg/m^2^) | 19.0 | (17.2, 21.3) | 18.7 | (16.8, 20.2) | 19.1 | (17.1, 21.5) | 17.7 | (16.4, 19.3) |
| Waist circumference (cm) | 71.0 | (66.5, 77.5) | 67.5 | (63.3, 71.8) | 70.3 | (66.9, 78.7) | 66.1 | (62.8, 70.1) |
| Hip circumference (cm) | 87.0 | (82.3, 92.4) | 86.1 | (82.0, 89.9) | 86.8 | (83.0, 93.0) | 85.9 | (81.8, 90.8) |
| WHR | 0.82 | (0.79, 0.85) | 0.78 | (0.75, 0.81) | 0.82 | (0.78, 0.85) | 0.77 | (0.74, 0.81) |
| Sum of skinfolds (mm) | 32.6 | (24.0, 52.3) | 57.3 | (42.3, 79.9) | 33.9 | (24.2, 49.1) | 49.4 | (38.7, 63.4) |
| DXA Fat percentage (%) | 13.0 | (8.7, 20.6) | 28.1 | (22.8, 32.4) | 12.8 | (9.8, 19.1) | 27.6 | (24.9, 30.3) |
| Fasting glucose (mg%) | 97.0 | (93.0, 101.0) | 93.0 | (89.0, 96.0) | 95.0 | (91.7, 99.0) | 91.0 | (89.7, 94.0) |
| **120-min glucose (mg%)** | 110.0 | (96.0, 126.2) | **114.0** | **(99.0, 128.0)** | 103.5 | (88.0, 123.0) | **103.0*** | **(97.0, 115.0)** |

Supplementary Table S4: Associations of ASAT and VAT at 18-years with cardiometabolic markers at 18-years

|  | **Men** | | | | | **Women** | | | | |
| --- | --- | --- | --- | --- | --- | --- | --- | --- | --- | --- |
|  | **ASAT (18-years)** | | **VAT (18-years)** | | **Diff. b/w coefficients** | **ASAT (18-years)** | | **VAT (18-years)** | | **Diff. b/w coefficients** |
| **Outcome at 18-years** | **β (SE)** | **p-value** | **β (SE)** | **p-value** | **Z (p-value)** | **β (SE)** | **p-value** | **β (SE)** | **p-value** | **Z (p-value)** |
| Fasting glucose | 0.020 (0.067) | 0.763 | 0.118 (0.067) | 0.079 | -1.03 (0.301) | 0.020 (0.066) | 0.764 | 0.089 (0.066) | 0.177 | -0.74 (0.457) |
| 120-minute glucose | 0.127 (0.067) | 0.060 | 0.061 (0.067) | 0.361 | 0.69 (0.488) | 0.115 (0.071) | 0.107 | -0.098 (0.071) | 0.169 | 2.12 (0.034) |
| Insulinogenic Index | 0.037 (0.068) | 0.590 | -0.054 (0.067) | 0.426 | 0.94 (0.345) | 0.013 (0.072) | 0.859 | -0.038 (0.071) | 0.597 | 0.50 (0.618) |
| Matsuda Index | -0.454 (0.057) | <0.001 | -0.132 (0.057) | 0.021 | -4.00 (<0.001) | -0.169 (0.068) | 0.013 | -0.054 (0.067) | 0.423 | -1.21 (0.224) |
| Disposition Index | -0.318 (0.062) | <0.001 | -0.117 (0.062) | 0.059 | -2.28 (0.023) | -0.181 (0.071) | 0.011 | 0.019 (0.070) | 0.791 | -2.00 (0.045) |
| Systolic BP | 0.188 (0.064) | 0.004 | 0.184 (0.064) | 0.004 | 0.04 (0.970) | 0.312 (0.063) | <0.001 | 0.021 (0.063) | 0.733 | 3.27 (0.001) |
| Diastolic BP | 0.257 (0.065) | <0.001 | 0.035 (0.065) | 0.586 | 2.42 (0.016) | 0.359 (0.062) | <0.001 | -0.010 (0.062) | 0.875 | 4.20 (<0.001) |
| Triglycerides | 0.322 (0.063) | <0.001 | 0.062 (0.063) | 0.330 | 2.93 (0.003) | 0.187 (0.065) | 0.004 | 0.049 (0.065) | 0.455 | 1.51 (0.132) |
| Total cholesterol | 0.292 (0.063) | <0.001 | 0.079 (0.063) | 0.215 | 2.38 (0.017) | 0.181 (0.066) | 0.006 | -0.040 (0.065) | 0.543 | 2.38 (0.017) |
| HDL-cholesterol | -0.074 (0.067) | 0.273 | -0.005 (0.067) | 0.945 | -0.73 (0.468) | -0.078 (0.065) | 0.232 | -0.160 (0.065) | 0.014 | 0.90 (0.369) |
| LDL-cholesterol | 0.378 (0.064) | <0.001 | -0.002 (0.063) | 0.979 | 4.24 (<0.001) | 0.214 (0.065) | 0.001 | 0.012 (0.065) | 0.854 | 2.20 (0.028) |
| TG:HDL-c | 0.289 (0.064) | <0.001 | 0.051 (0.064) | 0.426 | 2.63 (0.008) | 0.186 (0.064) | 0.004 | 0.114 (0.064) | 0.076 | 0.79 (0.428) |
| Leptin | 0.817 (0.036) | <0.001 | 0.056 (0.036) | 0.120 | 15.06 (<0.001) | 0.765 (0.044) | <0.001 | -0.035 (0.044) | 0.426 | 12.92 (<0.001) |
| Total Adiponectin | -0.303 (0.064) | <0.001 | -0.091 (0.063) | 0.152 | -2.35 (0.019) | -0.260 (0.065) | <0.001 | 0.011 (0.065) | 0.864 | -2.94 (0.003) |
| Total leukocyte count | 0.153 (0.066) | 0.021 | 0.050 (0.066) | 0.449 | 1.10 (0.270) | 0.168 (0.065) | 0.011 | 0.009 (0.065) | 0.895 | 1.73 (0.084) |
| C-reactive protein | 0.153 (0.068) | 0.025 | 0.033 (0.067) | 0.626 | 1.26 (0.209) | 0.160 (0.072) | 0.028 | -0.045 (0.072) | 0.531 | 2.01 (0.044) |

Values are β (SE) from multivariable linear regression models. Diff. b/w coefficients = difference between the ASAT and VAT β-coefficients within each sex, tested using a Wald’s test (Z statistic, and p-value shown in parentheses). ASAT, abdominal subcutaneous adipose tissue; VAT, visceral adipose tissue; SE, standard error; BP, blood pressure.

Supplementary Table S5: Associations of ASAT and VAT at 18-years with cardiometabolic markers at 24-years

|  | Men | | | | | Women | | | | |
| --- | --- | --- | --- | --- | --- | --- | --- | --- | --- | --- |
|  | ASAT (18-years) | | VAT (18-years) | | Diff. b/w coefficients | ASAT (18-years) | | VAT (18-years) | | Diff. b/w coefficients |
| Outcome at 24-years | β (SE) | p-value | β (SE) | p-value | Z (p-value) | β (SE) | p-value | β (SE) | p-value | Z (p-value) |
| Fasting glucose | 0.055 (0.071) | 0.444 | 0.089 (0.070) | 0.207 | -0.34 (0.731) | 0.041 (0.069) | 0.547 | 0.108 (0.069) | 0.122 | -0.68 (0.498) |
| 120-minute glucose | 0.141 (0.071) | 0.047 | 0.036 (0.070) | 0.604 | 1.05 (0.294) | 0.092 (0.069) | 0.188 | -0.003 (0.070) | 0.971 | 0.96 (0.339) |
| Insulinogenic Index | -0.199 (0.070) | 0.005 | -0.019 (0.070) | 0.787 | -1.82 (0.069) | -0.215 (0.068) | 0.002 | -0.051 (0.069) | 0.462 | -1.71 (0.088) |
| Matsuda Index | -0.462 (0.063) | <0.001 | -0.007 (0.063) | 0.905 | -5.09 (<0.001) | -0.262 (0.067) | <0.001 | -0.017 (0.068) | 0.803 | -2.57 (0.010) |
| Disposition Index | -0.452 (0.064) | <0.001 | -0.013 (0.063) | 0.835 | -4.87 (<0.001) | -0.310 (0.066) | <0.001 | -0.033 (0.067) | 0.621 | -2.93 (0.003) |
| Systolic BP | 0.237 (0.067) | <0.001 | 0.068 (0.067) | 0.308 | 1.78 (0.075) | 0.214 (0.067) | 0.002 | 0.034 (0.068) | 0.614 | 1.87 (0.061) |
| Diastolic BP | 0.290 (0.068) | <0.001 | -0.004 (0.067) | 0.951 | 3.09 (0.002) | 0.262 (0.067) | <0.001 | -0.024 (0.068) | 0.724 | 2.99 (0.003) |
| Triglycerides | 0.288 (0.067) | <0.001 | 0.025 (0.067) | 0.705 | 2.77 (0.006) | 0.190 (0.068) | 0.006 | -0.026 (0.069) | 0.710 | 2.22 (0.027) |
| Total cholesterol | 0.189 (0.069) | 0.007 | -0.044 (0.069) | 0.527 | 2.38 (0.017) | 0.100 (0.069) | 0.148 | -0.009 (0.070) | 0.900 | 1.11 (0.267) |
| HDL-cholesterol | -0.157 (0.069) | 0.024 | -0.048 (0.069) | 0.489 | -1.12 (0.263) | -0.263 (0.066) | <0.001 | -0.080 (0.067) | 0.234 | -1.95 (0.051) |
| LDL-cholesterol | 0.166 (0.070) | 0.018 | -0.056 (0.069) | 0.420 | 2.26 (0.024) | 0.177 (0.068) | 0.010 | 0.028 (0.069) | 0.690 | 1.54 (0.123) |
| TG:HDL-c | 0.300 (0.067) | <0.001 | 0.038 (0.066) | 0.565 | 2.78 (0.005) | 0.270 (0.067) | <0.001 | 0.009 (0.068) | 0.892 | 2.74 (0.006) |
| Total leukocyte count | 0.320 (0.068) | <0.001 | -0.106 (0.067) | 0.118 | 4.44 (<0.001) | 0.249 (0.068) | <0.001 | -0.009 (0.068) | 0.900 | 2.68 (0.007) |
| C-reactive protein | 0.445 (0.065) | <0.001 | -0.115 (0.064) | 0.074 | 6.14 (<0.001) | 0.423 (0.064) | <0.001 | -0.148 (0.064) | 0.022 | 6.30 (<0.001) |

Values are β (SE) from multivariable linear regression models. Diff. b/w coefficients = difference between the ASAT and VAT β-coefficients within each sex, tested using a Wald’s test (Z statistic, and p-value shown in parentheses). ASAT, abdominal subcutaneous adipose tissue; VAT, visceral adipose tissue; SE, standard error; BP, blood pressure. Leptin and Total Adiponectin were not assessed at the 24-year follow-up.

Supplementary Table S6: Associations of MRI ASAT and VAT with change in outcome (Delta) between 18- and 24-years

|  | **Men** | | | | **Women** | | | |
| --- | --- | --- | --- | --- | --- | --- | --- | --- |
|  | **ASAT (18-years)** | | **VAT (18-years)** | | **ASAT (18-years)** | | **VAT (18-years)** | |
| **Delta Outcome (24-years – 18-years)** | **β (SE)** | **p-value** | **β (SE)** | **p-value** | **β (SE)** | **p-value** | **β (SE)** | **p-value** |
| Fasting glucose | 0.110 (0.086) | 0.204 | -0.064 (0.084) | 0.441 | 0.130 (0.076) | 0.089 | 0.035 (0.076) | 0.645 |
| 120_minute_glucose | 0.036 (0.096) | 0.708 | -0.010 (0.093) | 0.913 | 0.046 (0.106) | 0.666 | -0.006 (0.105) | 0.953 |
| Insulinogenic Index | -0.253 (0.103) | **0.015** | -0.008 (0.099) | 0.933 | -0.142 (0.098) | 0.149 | -0.074 (0.098) | 0.450 |
| Matsuda Index | -0.040 (0.080) | 0.613 | 0.164 (0.077) | **0.034** | -0.214 (0.098) | **0.030** | 0.139 (0.097) | 0.152 |
| Disposition Index | -0.169 (0.091) | 0.064 | 0.116 (0.087) | 0.187 | -0.180 (0.107) | 0.094 | 0.004 (0.107) | 0.972 |
| Systolic BP | 0.001 (0.073) | 0.992 | -0.046 (0.071) | 0.516 | -0.085 (0.082) | 0.296 | 0.035 (0.082) | 0.667 |
| Diastolic BP | -0.072 (0.087) | 0.408 | 0.054 (0.084) | 0.522 | -0.076 (0.086) | 0.378 | -0.034 (0.086) | 0.692 |
| Triglycerides | 0.018 (0.072) | 0.808 | -0.094 (0.070) | 0.182 | 0.088 (0.089) | 0.322 | -0.094 (0.089) | 0.289 |
| Total cholesterol | -0.054 (0.072) | 0.450 | -0.111 (0.069) | 0.113 | -0.057 (0.077) | 0.461 | 0.047 (0.077) | 0.543 |
| HDL cholesterol | -0.132 (0.075) | 0.080 | 0.069 (0.073) | 0.343 | -0.256 (0.082) | **0.002** | 0.188 (0.082) | **0.023** |
| LDL cholesterol | -0.206 (0.067) | **0.002** | -0.036 (0.064) | 0.575 | 0.008 (0.076) | 0.916 | -0.006 (0.075) | 0.938 |
| TG:HDL-c | 0.074 (0.071) | 0.302 | -0.103 (0.069) | 0.137 | 0.187 (0.091) | **0.041** | -0.168 (0.090) | 0.063 |
| C-reactive protein | 0.232 (0.113) | **0.042** | -0.156 (0.108) | 0.149 | 0.215 (0.109) | **0.050** | -0.060 (0.107) | 0.576 |
| Total leukocyte count | 0.217 (0.074) | **0.003** | -0.203 (0.071) | **0.005** | 0.036 (0.087) | 0.675 | -0.039 (0.087) | 0.658 |

Values are β (SE) from multivariable linear regression models. ASAT, abdominal subcutaneous adipose tissue; VAT, visceral adipose tissue; SE, standard error; BP, blood pressure. Leptin and Total Adiponectin were not assessed at the 24-year follow-up.

Supplementary Table S7: Associations of ASAT and VAT at 18-years with cardiometabolic markers at 18- and 24years with an additional adjustment for height at 18-years

|  | Men | | | | | Women | | | | |
| --- | --- | --- | --- | --- | --- | --- | --- | --- | --- | --- |
|  | ASAT (18-years) | | VAT (18-years) | | Diff. b/w coefficients | ASAT (18-years) | | VAT (18-years) | | Diff. b/w coefficients |
| Outcome Variable | β (SE) | p-value | β (SE) | p-value | Z (p-value) | β (SE) | p-value | β (SE) | p-value | Z (p-value) |
| Outcomes at 18-Years | | | | | | | | | | |
| Fasting glucose | 0.023 (0.067) | 0.728 | 0.121 (0.067) | 0.071 | -1.04 (0.300) | 0.013 (0.067) | 0.848 | 0.094 (0.066) | 0.158 | -0.86 (0.389) |
| 120-minute glucose | 0.138 (0.067) | 0.040 | 0.072 (0.066) | 0.280 | 0.70 (0.481) | 0.087 (0.071) | 0.219 | -0.079 (0.070) | 0.259 | 1.67 (0.095) |
| Insulinogenic Index | 0.034 (0.068) | 0.622 | -0.058 (0.068) | 0.394 | 0.95 (0.341) | 0.040 (0.072) | 0.575 | -0.056 (0.071) | 0.433 | 0.95 (0.342) |
| Matsuda Index | -0.467 (0.056) | <0.001 | -0.147 (0.056) | 0.009 | -4.03 (<0.001) | -0.164 (0.069) | 0.018 | -0.057 (0.067) | 0.397 | -1.11 (0.266) |
| Disposition Index | -0.328 (0.062) | <0.001 | -0.130 (0.062) | 0.036 | -2.26 (0.024) | -0.156 (0.071) | 0.029 | 0.003 (0.070) | 0.968 | -1.60 (0.110) |
| Systolic BP | 0.188 (0.064) | 0.004 | 0.184 (0.064) | 0.004 | 0.04 (0.970) | 0.325 (0.063) | <0.001 | 0.013 (0.063) | 0.831 | 3.48 (<0.001) |
| Diastolic BP | 0.263 (0.065) | <0.001 | 0.043 (0.065) | 0.509 | 2.40 (0.017) | 0.347 (0.063) | <0.001 | -0.002 (0.062) | 0.976 | 3.95 (<0.001) |
| Triglycerides | 0.330 (0.063) | <0.001 | 0.070 (0.063) | 0.265 | 2.91 (0.004) | 0.164 (0.065) | 0.012 | 0.063 (0.065) | 0.329 | 1.10 (0.270) |
| Total cholesterol | 0.303 (0.063) | <0.001 | 0.092 (0.063) | 0.144 | 2.36 (0.018) | 0.160 (0.066) | 0.015 | -0.026 (0.065) | 0.686 | 2.02 (0.044) |
| HDL-cholesterol | -0.076 (0.067) | 0.258 | -0.008 (0.068) | 0.907 | -0.72 (0.473) | -0.070 (0.066) | 0.289 | -0.166 (0.065) | 0.012 | 1.04 (0.300) |
| LDL-cholesterol | 0.390 (0.063) | <0.001 | 0.014 (0.062) | 0.825 | 4.25 (<0.001) | 0.190 (0.065) | 0.004 | 0.027 (0.064) | 0.670 | 1.78 (0.076) |
| TG:HDL-c | 0.296 (0.064) | <0.001 | 0.060 (0.064) | 0.353 | 2.61 (0.009) | 0.164 (0.064) | 0.011 | 0.128 (0.064) | 0.045 | 0.40 (0.692) |
| Leptin | 0.819 (0.036) | <0.001 | 0.059 (0.036) | 0.103 | 15.01 (<0.001) | 0.764 (0.044) | <0.001 | -0.034 (0.044) | 0.436 | 12.79 (<0.001) |
| Total Adiponectin | -0.308 (0.064) | <0.001 | -0.096 (0.064) | 0.133 | -2.34 (0.019) | -0.257 (0.066) | <0.001 | 0.009 (0.065) | 0.892 | -2.85 (0.004) |
| Total leukocyte count | 0.155 (0.066) | 0.020 | 0.053 (0.066) | 0.428 | 1.10 (0.274) | 0.143 (0.065) | 0.030 | 0.025 (0.065) | 0.698 | 1.28 (0.202) |
| C-reactive protein | 0.165 (0.068) | 0.016 | 0.046 (0.067) | 0.494 | 1.24 (0.214) | 0.150 (0.073) | 0.040 | -0.038 (0.072) | 0.596 | 1.84 (0.065) |
| Outcomes at 24-Years | | | | | | | | | | |
| Fasting glucose | 0.134 (0.076) | 0.079 | 0.057 (0.074) | 0.446 | 0.73 (0.466) | 0.120 (0.083) | 0.149 | 0.089 (0.082) | 0.278 | 0.26 (0.793) |
| 120-minute glucose | 0.128 (0.076) | 0.094 | 0.080 (0.074) | 0.284 | 0.45 (0.651) | 0.090 (0.083) | 0.281 | -0.069 (0.082) | 0.402 | 1.36 (0.174) |
| Insulinogenic Index | -0.158 (0.076) | 0.039 | -0.064 (0.074) | 0.393 | -0.89 (0.375) | -0.196 (0.083) | 0.020 | -0.017 (0.083) | 0.841 | -1.52 (0.128) |
| Matsuda Index | -0.486 (0.068) | <0.001 | 0.003 (0.066) | 0.967 | -5.18 (<0.001) | -0.406 (0.079) | <0.001 | 0.086 (0.078) | 0.269 | -4.45 (<0.001) |
| Disposition Index | -0.451 (0.068) | <0.001 | -0.027 (0.067) | 0.682 | -4.44 (<0.001) | -0.406 (0.079) | <0.001 | 0.066 (0.079) | 0.405 | -4.24 (<0.001) |
| Systolic BP | 0.228 (0.074) | 0.002 | 0.072 (0.073) | 0.320 | 1.50 (0.133) | 0.295 (0.080) | <0.001 | 0.010 (0.079) | 0.900 | 2.54 (0.011) |
| Diastolic BP | 0.288 (0.075) | <0.001 | 0.012 (0.073) | 0.869 | 2.64 (0.008) | 0.331 (0.079) | <0.001 | -0.090 (0.079) | 0.254 | 3.77 (<0.001) |
| Triglycerides | 0.343 (0.073) | <0.001 | 0.006 (0.071) | 0.933 | 3.30 (<0.001) | 0.240 (0.083) | 0.004 | -0.126 (0.082) | 0.125 | 3.15 (0.002) |
| Total cholesterol | 0.215 (0.076) | 0.005 | -0.048 (0.074) | 0.517 | 2.47 (0.013) | 0.140 (0.084) | 0.097 | -0.101 (0.083) | 0.227 | 2.04 (0.041) |
| HDL-cholesterol | -0.201 (0.076) | 0.009 | 0.015 (0.075) | 0.844 | -2.02 (0.043) | -0.332 (0.079) | <0.001 | -0.036 (0.078) | 0.646 | -2.66 (0.008) |
| LDL-cholesterol | 0.187 (0.076) | 0.015 | -0.070 (0.075) | 0.346 | 2.41 (0.016) | 0.249 (0.082) | 0.003 | -0.076 (0.082) | 0.356 | 2.80 (0.005) |
| TG:HDL-c | 0.363 (0.073) | <0.001 | -0.000 (0.071) | 0.999 | 3.58 (<0.001) | 0.343 (0.081) | <0.001 | -0.090 (0.080) | 0.260 | 3.82 (<0.001) |
| Total leukocyte count | 0.351 (0.074) | <0.001 | -0.143 (0.073) | 0.050 | 4.76 (<0.001) | 0.215 (0.082) | 0.009 | -0.058 (0.082) | 0.479 | 2.36 (0.018) |
| C-reactive protein | 0.462 (0.071) | <0.001 | -0.139 (0.069) | 0.046 | 6.06 (<0.001) | 0.521 (0.075) | <0.001 | -0.202 (0.073) | 0.006 | 6.93 (<0.001) |

Values are β (SE) from multivariable linear regression models. Diff. b/w coefficients = difference between the ASAT and VAT β-coefficients within each sex, tested using a Wald’s test (Z statistic, and p-value shown in parentheses). ASAT, abdominal subcutaneous adipose tissue; VAT, visceral adipose tissue; SE, standard error; BP, blood pressure. Leptin and Total Adiponectin were not assessed at the 24-year follow-up.

Supplementary Table S8: Associations of ASAT and VAT at 18-years with cardiometabolic markers at 18- and 24years with an additional adjustment for BMI at 18-years

|  | Men | | | | | Women | | | | |
| --- | --- | --- | --- | --- | --- | --- | --- | --- | --- | --- |
|  | ASAT (18-years) | | VAT (18-years) | | Diff. b/w coefficients | ASAT (18-years) | | VAT (18-years) | | Diff. b/w coefficients |
| Outcome | β (SE) | p-value | β (SE) | p-value | Z (p-value) | β (SE) | p-value | β (SE) | p-value | Z (p-value) |
| Outcomes assessed at 18-years | | | | | | | | | | |
| Fasting glucose | -0.059 (0.123) | 0.634 | 0.113 (0.067) | 0.094 | -1.22 (0.222) | 0.096 (0.124) | 0.439 | 0.100 (0.068) | 0.140 | -0.03 (0.977) |
| 120-minute glucose | 0.407 (0.122) | <0.001 | 0.078 (0.066) | 0.239 | 2.37 (0.018) | 0.302 (0.130) | 0.021 | -0.071 (0.072) | 0.327 | 2.52 (0.012) |
| Insulinogenic Index | 0.188 (0.125) | 0.136 | -0.045 (0.068) | 0.509 | 1.63 (0.103) | -0.022 (0.131) | 0.867 | -0.043 (0.073) | 0.559 | 0.14 (0.890) |
| Matsuda Index | -0.463 (0.105) | <0.001 | -0.132 (0.057) | 0.021 | -2.76 (0.006) | -0.062 (0.124) | 0.618 | -0.037 (0.069) | 0.596 | -0.18 (0.859) |
| Disposition Index | -0.169 (0.115) | 0.142 | -0.109 (0.062) | 0.081 | -0.46 (0.642) | -0.163 (0.129) | 0.209 | 0.021 (0.072) | 0.770 | -1.24 (0.213) |
| Systolic BP | 0.124 (0.118) | 0.293 | 0.180 (0.064) | 0.005 | -0.42 (0.675) | 0.021 (0.117) | 0.858 | -0.020 (0.064) | 0.753 | 0.31 (0.758) |
| Diastolic BP | 0.346 (0.120) | 0.004 | 0.041 (0.065) | 0.531 | 2.24 (0.025) | 0.386 (0.117) | 0.001 | -0.006 (0.064) | 0.926 | 2.95 (0.003) |
| Triglycerides | 0.102 (0.115) | 0.377 | 0.048 (0.063) | 0.448 | 0.41 (0.680) | 0.046 (0.122) | 0.706 | 0.028 (0.066) | 0.669 | 0.13 (0.899) |
| Total cholesterol | 0.152 (0.117) | 0.193 | 0.070 (0.064) | 0.271 | 0.62 (0.537) | 0.059 (0.123) | 0.632 | -0.057 (0.067) | 0.394 | 0.83 (0.406) |
| HDL-cholesterol | 0.053 (0.124) | 0.671 | 0.003 (0.068) | 0.963 | 0.35 (0.726) | 0.054 (0.122) | 0.658 | -0.142 (0.067) | 0.034 | 1.41 (0.159) |
| LDL-cholesterol | 0.220 (0.115) | 0.058 | -0.011 (0.063) | 0.856 | 1.76 (0.079) | 0.050 (0.121) | 0.679 | -0.011 (0.066) | 0.863 | 0.45 (0.655) |
| TG:HDL-c | 0.058 (0.117) | 0.619 | 0.037 (0.064) | 0.565 | 0.16 (0.872) | 0.011 (0.120) | 0.924 | 0.089 (0.066) | 0.175 | -0.57 (0.570) |
| Leptin | 0.977 (0.065) | <0.001 | 0.066 (0.035) | 0.063 | 12.29 (<0.001) | 0.535 (0.081) | <0.001 | -0.068 (0.044) | 0.125 | 6.56 (<0.001) |
| Total Adiponectin | -0.090 (0.116) | 0.436 | -0.078 (0.063) | 0.221 | -0.10 (0.923) | -0.107 (0.121) | 0.375 | 0.032 (0.066) | 0.630 | -1.01 (0.312) |
| Total leukocyte count | 0.097 (0.122) | 0.427 | 0.046 (0.067) | 0.486 | 0.36 (0.715) | 0.199 (0.123) | 0.107 | 0.013 (0.067) | 0.847 | 1.33 (0.185) |
| C-reactive protein | -0.034 (0.123) | 0.781 | 0.022 (0.067) | 0.747 | -0.40 (0.690) | 0.152 (0.134) | 0.260 | -0.046 (0.073) | 0.531 | 1.29 (0.196) |
| Outcomes assessed at 24-years | | | | | | | | | | |
| Fasting glucose | 0.114 (0.139) | 0.414 | 0.069 (0.075) | 0.357 | 0.28 (0.779) | 0.233 (0.155) | 0.133 | 0.110 (0.083) | 0.189 | 0.70 (0.482) |
| 120-minute glucose | 0.388 (0.137) | 0.005 | 0.109 (0.074) | 0.144 | 1.79 (0.073) | 0.222 (0.157) | 0.159 | -0.081 (0.085) | 0.342 | 1.70 (0.090) |
| Insulinogenic Index | -0.309 (0.138) | 0.026 | -0.087 (0.075) | 0.245 | -1.41 (0.157) | -0.169 (0.155) | 0.278 | -0.025 (0.085) | 0.770 | -0.81 (0.415) |
| Matsuda Index | -0.624 (0.122) | <0.001 | -0.017 (0.066) | 0.795 | -4.36 (<0.001) | -0.502 (0.148) | <0.001 | 0.046 (0.080) | 0.562 | -3.25 (0.001) |
| Disposition Index | -0.628 (0.123) | <0.001 | -0.054 (0.067) | 0.422 | -4.09 (<0.001) | -0.464 (0.148) | 0.002 | 0.031 (0.081) | 0.700 | -2.94 (0.003) |
| Systolic BP | 0.229 (0.135) | 0.091 | 0.069 (0.073) | 0.343 | 1.04 (0.298) | 0.201 (0.150) | 0.180 | 0.011 (0.081) | 0.891 | 1.12 (0.264) |
| Diastolic BP | 0.481 (0.135) | <0.001 | 0.031 (0.073) | 0.673 | 2.95 (0.003) | 0.343 (0.149) | 0.023 | -0.103 (0.081) | 0.203 | 2.63 (0.009) |
| Triglycerides | 0.412 (0.132) | 0.002 | 0.016 (0.072) | 0.822 | 2.63 (0.009) | 0.202 (0.155) | 0.194 | -0.128 (0.083) | 0.126 | 1.88 (0.060) |
| Total cholesterol | 0.050 (0.138) | 0.716 | -0.074 (0.075) | 0.322 | 0.79 (0.428) | 0.111 (0.157) | 0.482 | -0.113 (0.085) | 0.185 | 1.25 (0.211) |
| HDL-cholesterol | -0.221 (0.139) | 0.112 | 0.013 (0.075) | 0.861 | -1.49 (0.137) | -0.331 (0.148) | 0.027 | -0.028 (0.080) | 0.727 | -1.80 (0.071) |
| LDL-cholesterol | -0.032 (0.138) | 0.815 | -0.104 (0.075) | 0.165 | 0.46 (0.647) | 0.220 (0.155) | 0.157 | -0.092 (0.083) | 0.272 | 1.77 (0.076) |
| TG:HDL-c | 0.429 (0.132) | 0.001 | 0.009 (0.071) | 0.898 | 2.81 (0.005) | 0.308 (0.151) | 0.042 | -0.096 (0.081) | 0.239 | 2.36 (0.018) |
| Total leukocyte count | 0.367 (0.135) | 0.007 | -0.146 (0.073) | 0.047 | 3.34 (<0.001) | 0.351 (0.156) | 0.025 | -0.069 (0.084) | 0.412 | 2.38 (0.017) |
| C-reactive protein | 0.514 (0.129) | <0.001 | -0.133 (0.070) | 0.059 | 4.41 (<0.001) | 0.372 (0.139) | 0.008 | -0.221 (0.074) | 0.003 | 3.77 (<0.001) |

Values are β (SE) from multivariable linear regression models. Diff. b/w coefficients = difference between the ASAT and VAT β-coefficients within each sex, tested using a Wald’s test (Z statistic, and p-value shown in parentheses). ASAT, abdominal subcutaneous adipose tissue; VAT, visceral adipose tissue; SE, standard error; BP, blood pressure. Leptin and Total Adiponectin were not assessed at the 24-year follow-up.

Supplementary Table S9: Associations of ASAT and VAT at 18-years with cardiometabolic markers at 18- and 24years with an additional adjustment for waist circumference at 18-years

|  | Men | | | | | Women | | | | |
| --- | --- | --- | --- | --- | --- | --- | --- | --- | --- | --- |
|  | ASAT (18-years) | | VAT (18-years) | | Diff. b/w coefficients | ASAT (18-years) | | VAT (18-years) | | Diff. b/w coefficients |
| Outcome | β (SE) | p-value | β (SE) | p-value | Z (p-value) | β (SE) | p-value | β (SE) | p-value | Z (p-value) |
| Outcomes assessed at 18-years | | | | | | | | | | |
| Fasting glucose | -0.068 (0.131) | 0.604 | 0.105 (0.069) | 0.127 | -1.17 (0.242) | 0.026 (0.112) | 0.818 | 0.091 (0.069) | 0.191 | -0.49 (0.624) |
| 120-minute glucose | 0.324 (0.133) | 0.015 | 0.088 (0.068) | 0.199 | 1.58 (0.114) | 0.312 (0.115) | 0.007 | -0.046 (0.074) | 0.532 | 2.62 (0.009) |
| Insulinogenic Index | 0.313 (0.135) | 0.021 | -0.016 (0.069) | 0.816 | 2.18 (0.029) | -0.119 (0.117) | 0.311 | -0.072 (0.075) | 0.339 | -0.34 (0.735) |
| Matsuda Index | -0.469 (0.113) | <0.001 | -0.134 (0.059) | 0.023 | -2.64 (0.008) | -0.106 (0.112) | 0.344 | -0.038 (0.071) | 0.591 | -0.51 (0.607) |
| Disposition Index | -0.106 (0.124) | 0.396 | -0.088 (0.063) | 0.164 | -0.12 (0.902) | -0.254 (0.115) | 0.029 | -0.000 (0.074) | 0.996 | -1.85 (0.064) |
| Systolic BP | 0.051 (0.126) | 0.686 | 0.166 (0.066) | 0.012 | -0.81 (0.419) | 0.213 (0.107) | 0.048 | -0.001 (0.066) | 0.988 | 1.70 (0.089) |
| Diastolic BP | 0.244 (0.128) | 0.057 | 0.034 (0.067) | 0.615 | 1.46 (0.145) | 0.363 (0.106) | <0.001 | -0.009 (0.065) | 0.890 | 2.99 (0.003) |
| Triglycerides | 0.111 (0.124) | 0.370 | 0.033 (0.064) | 0.612 | 0.56 (0.575) | 0.316 (0.110) | 0.004 | 0.078 (0.068) | 0.252 | 1.84 (0.065) |
| Total cholesterol | 0.310 (0.125) | 0.014 | 0.081 (0.065) | 0.214 | 1.62 (0.105) | 0.243 (0.111) | 0.030 | -0.026 (0.069) | 0.708 | 2.06 (0.040) |
| HDL-cholesterol | 0.052 (0.133) | 0.697 | 0.012 (0.069) | 0.857 | 0.26 (0.793) | 0.105 (0.110) | 0.342 | -0.119 (0.068) | 0.079 | 1.74 (0.083) |
| LDL-cholesterol | 0.375 (0.124) | 0.003 | -0.002 (0.065) | 0.974 | 2.70 (0.007) | 0.189 (0.110) | 0.088 | 0.006 (0.068) | 0.925 | 1.41 (0.158) |
| TG:HDL-c | 0.066 (0.125) | 0.600 | 0.021 (0.065) | 0.753 | 0.32 (0.749) | 0.204 (0.109) | 0.063 | 0.118 (0.067) | 0.080 | 0.67 (0.503) |
| Leptin | 0.878 (0.070) | <0.001 | 0.064 (0.037) | 0.080 | 10.27 (<0.001) | 0.573 (0.073) | <0.001 | -0.078 (0.045) | 0.083 | 7.58 (<0.001) |
| Total Adiponectin | -0.169 (0.125) | 0.178 | -0.072 (0.065) | 0.268 | -0.69 (0.493) | -0.058 (0.109) | 0.595 | 0.057 (0.068) | 0.398 | -0.90 (0.369) |
| Total leukocyte count | -0.061 (0.129) | 0.635 | 0.020 (0.068) | 0.773 | -0.55 (0.579) | 0.035 (0.111) | 0.756 | -0.022 (0.068) | 0.752 | 0.43 (0.667) |
| C-reactive protein | 0.128 (0.135) | 0.345 | 0.029 (0.069) | 0.673 | 0.65 (0.516) | 0.160 (0.118) | 0.175 | -0.045 (0.076) | 0.554 | 1.47 (0.143) |
| Outcomes assessed at 24-years | | | | | | | | | | |
| Fasting glucose | -0.067 (0.144) | 0.641 | 0.041 (0.076) | 0.590 | -0.67 (0.506) | 0.138 (0.138) | 0.321 | 0.099 (0.086) | 0.252 | 0.24 (0.811) |
| 120-minute glucose | 0.419 (0.143) | 0.004 | 0.125 (0.075) | 0.099 | 1.83 (0.068) | 0.363 (0.139) | 0.010 | -0.036 (0.086) | 0.676 | 2.45 (0.014) |
| Insulinogenic Index | -0.203 (0.144) | 0.160 | -0.078 (0.076) | 0.310 | -0.77 (0.440) | 0.036 (0.137) | 0.792 | 0.027 (0.086) | 0.752 | 0.06 (0.956) |
| Matsuda Index | -0.667 (0.127) | <0.001 | -0.030 (0.067) | 0.660 | -4.42 (<0.001) | -0.292 (0.133) | 0.029 | 0.085 (0.082) | 0.303 | -2.41 (0.016) |
| Disposition Index | -0.613 (0.129) | <0.001 | -0.059 (0.068) | 0.387 | -3.80 (<0.001) | -0.215 (0.132) | 0.105 | 0.085 (0.083) | 0.307 | -1.92 (0.054) |
| Systolic BP | 0.094 (0.141) | 0.506 | 0.049 (0.074) | 0.510 | 0.28 (0.778) | 0.213 (0.134) | 0.114 | 0.006 (0.084) | 0.945 | 1.31 (0.191) |
| Diastolic BP | 0.433 (0.141) | 0.002 | 0.032 (0.074) | 0.670 | 2.52 (0.012) | 0.406 (0.133) | 0.003 | -0.087 (0.083) | 0.296 | 3.14 (0.002) |
| Triglycerides | 0.324 (0.138) | 0.020 | 0.006 (0.073) | 0.937 | 2.03 (0.042) | 0.191 (0.138) | 0.170 | -0.135 (0.086) | 0.118 | 2.00 (0.046) |
| Total cholesterol | 0.237 (0.144) | 0.102 | -0.053 (0.076) | 0.488 | 1.78 (0.076) | 0.244 (0.140) | 0.083 | -0.083 (0.087) | 0.339 | 1.99 (0.047) |
| HDL-cholesterol | -0.054 (0.144) | 0.709 | 0.038 (0.076) | 0.622 | -0.56 (0.575) | -0.101 (0.130) | 0.440 | 0.030 (0.081) | 0.707 | -0.86 (0.392) |
| LDL-cholesterol | 0.187 (0.145) | 0.199 | -0.080 (0.077) | 0.295 | 1.63 (0.103) | 0.301 (0.138) | 0.030 | -0.076 (0.086) | 0.376 | 2.32 (0.020) |
| TG:HDL-c | 0.295 (0.137) | 0.033 | -0.008 (0.073) | 0.908 | 1.95 (0.051) | 0.199 (0.134) | 0.140 | -0.127 (0.083) | 0.129 | 2.06 (0.039) |
| Total leukocyte count | 0.205 (0.140) | 0.145 | -0.170 (0.074) | 0.023 | 2.36 (0.018) | 0.229 (0.139) | 0.100 | -0.087 (0.087) | 0.315 | 1.94 (0.053) |
| C-reactive protein | 0.446 (0.135) | 0.001 | -0.141 (0.071) | 0.049 | 3.86 (<0.001) | 0.607 (0.124) | <0.001 | -0.177 (0.077) | 0.022 | 5.38 (<0.001) |

Values are β (SE) from multivariable linear regression models, additionally adjusted for waist circumference. Diff. b/w coefficients = difference between the ASAT and VAT β-coefficients within each sex, tested using a Wald’s test (Z statistic, and p-value shown in parentheses). ASAT, abdominal subcutaneous adipose tissue; VAT, visceral adipose tissue; SE, standard error; BP, blood pressure. Leptin and Total Adiponectin were not assessed at the 24-year follow-up.

Supplementary Table S10: Associations of ASAT and VAT at 18-years with cardiometabolic markers at 18- and 24years with an additional adjustment for birthweight

|  | Men | | | | | Women | | | | |
| --- | --- | --- | --- | --- | --- | --- | --- | --- | --- | --- |
|  | ASAT (18-years) | | VAT (18-years) | | Diff. b/w coefficients | ASAT (18-years) | | VAT (18-years) | | Diff. b/w coefficients |
| Outcome | β (SE) | p-value | β (SE) | p-value | Z (p-value) | β (SE) | p-value | β (SE) | p-value | Z (p-value) |
| Outcomes assessed at 18-years | | | | | | | | | | |
| Fasting glucose | 0.016 (0.068) | 0.820 | 0.102 (0.068) | 0.135 | -0.90 (0.370) | 0.029 (0.070) | 0.682 | 0.108 (0.069) | 0.119 | -0.81 (0.419) |
| 120-minute glucose | 0.125 (0.069) | 0.071 | 0.060 (0.068) | 0.382 | 0.68 (0.499) | 0.138 (0.074) | 0.064 | -0.087 (0.073) | 0.233 | 2.17 (0.030) |
| Insulinogenic Index | 0.036 (0.070) | 0.610 | -0.035 (0.069) | 0.609 | 0.72 (0.469) | -0.001 (0.075) | 0.984 | -0.041 (0.074) | 0.578 | 0.38 (0.707) |
| Matsuda Index | -0.471 (0.058) | <0.001 | -0.137 (0.058) | 0.018 | -4.08 (<0.001) | -0.179 (0.072) | 0.013 | -0.069 (0.070) | 0.327 | -1.10 (0.272) |
| Disposition Index | -0.329 (0.064) | <0.001 | -0.107 (0.063) | 0.091 | -2.46 (0.014) | -0.204 (0.074) | 0.006 | 0.008 (0.072) | 0.908 | -2.05 (0.040) |
| Systolic BP | 0.196 (0.067) | 0.004 | 0.179 (0.066) | 0.007 | 0.18 (0.858) | 0.331 (0.066) | <0.001 | 0.028 (0.065) | 0.663 | 3.29 (0.001) |
| Diastolic BP | 0.287 (0.067) | <0.001 | 0.025 (0.066) | 0.709 | 2.79 (0.005) | 0.377 (0.064) | <0.001 | -0.003 (0.064) | 0.968 | 4.19 (<0.001) |
| Triglycerides | 0.331 (0.064) | <0.001 | 0.052 (0.064) | 0.417 | 3.07 (0.002) | 0.218 (0.067) | 0.001 | 0.069 (0.066) | 0.298 | 1.60 (0.110) |
| Total cholesterol | 0.280 (0.065) | <0.001 | 0.080 (0.065) | 0.219 | 2.16 (0.030) | 0.196 (0.069) | 0.005 | -0.035 (0.068) | 0.609 | 2.39 (0.017) |
| HDL-cholesterol | -0.072 (0.070) | 0.300 | -0.009 (0.070) | 0.893 | -0.64 (0.522) | -0.068 (0.067) | 0.309 | -0.175 (0.066) | 0.009 | 1.14 (0.256) |
| LDL-cholesterol | 0.363 (0.065) | <0.001 | 0.008 (0.064) | 0.896 | 3.90 (<0.001) | 0.221 (0.068) | 0.001 | 0.019 (0.068) | 0.775 | 2.10 (0.036) |
| TG:HDL-c | 0.296 (0.065) | <0.001 | 0.046 (0.065) | 0.485 | 2.70 (0.007) | 0.207 (0.066) | 0.002 | 0.137 (0.065) | 0.035 | 0.76 (0.448) |
| Leptin | 0.809 (0.037) | <0.001 | 0.070 (0.036) | 0.057 | 14.29 (<0.001) | 0.770 (0.045) | <0.001 | -0.048 (0.045) | 0.281 | 12.83 (<0.001) |
| Total Adiponectin | -0.307 (0.066) | <0.001 | -0.090 (0.065) | 0.168 | -2.35 (0.019) | -0.254 (0.068) | <0.001 | 0.004 (0.067) | 0.955 | -2.72 (0.007) |
| Total leukocyte count | 0.186 (0.067) | 0.006 | 0.039 (0.067) | 0.558 | 1.55 (0.120) | 0.171 (0.067) | 0.011 | 0.036 (0.066) | 0.580 | 1.43 (0.151) |
| C-reactive protein | 0.178 (0.070) | 0.011 | 0.033 (0.068) | 0.633 | 1.49 (0.136) | 0.157 (0.076) | 0.041 | -0.042 (0.075) | 0.570 | 1.87 (0.062) |
| Outcomes assessed at 24-years | | | | | | | | | | |
| Fasting glucose | 0.133 (0.078) | 0.091 | 0.057 (0.076) | 0.452 | 0.69 (0.487) | 0.148 (0.081) | 0.068 | 0.107 (0.079) | 0.181 | 0.37 (0.712) |
| 120-minute glucose | 0.130 (0.078) | 0.095 | 0.073 (0.075) | 0.337 | 0.53 (0.593) | 0.148 (0.086) | 0.088 | -0.110 (0.085) | 0.194 | 2.14 (0.032) |
| Insulinogenic Index | -0.149 (0.078) | 0.058 | -0.067 (0.076) | 0.375 | -0.75 (0.453) | -0.191 (0.084) | 0.024 | -0.042 (0.083) | 0.609 | -1.26 (0.207) |
| Matsuda Index | -0.496 (0.068) | <0.001 | 0.003 (0.066) | 0.958 | -5.24 (<0.001) | -0.388 (0.079) | <0.001 | 0.066 (0.078) | 0.400 | -4.09 (<0.001) |
| Disposition Index | -0.455 (0.070) | <0.001 | -0.028 (0.067) | 0.674 | -4.40 (<0.001) | -0.391 (0.078) | <0.001 | 0.038 (0.077) | 0.618 | -3.91 (<0.001) |
| Systolic BP | 0.233 (0.076) | 0.002 | 0.053 (0.074) | 0.471 | 1.70 (0.089) | 0.300 (0.076) | <0.001 | -0.003 (0.075) | 0.971 | 2.83 (0.005) |
| Diastolic BP | 0.287 (0.076) | <0.001 | -0.008 (0.074) | 0.918 | 2.79 (0.005) | 0.358 (0.078) | <0.001 | -0.109 (0.078) | 0.162 | 4.25 (<0.001) |
| Triglycerides | 0.331 (0.073) | <0.001 | 0.005 (0.071) | 0.939 | 3.20 (0.001) | 0.268 (0.080) | 0.001 | -0.114 (0.079) | 0.149 | 3.39 (<0.001) |
| Total cholesterol | 0.200 (0.078) | 0.011 | -0.042 (0.076) | 0.581 | 2.22 (0.026) | 0.167 (0.087) | 0.055 | -0.104 (0.085) | 0.223 | 2.23 (0.026) |
| HDL-cholesterol | -0.178 (0.078) | 0.023 | 0.003 (0.075) | 0.971 | -1.67 (0.095) | -0.356 (0.081) | <0.001 | -0.016 (0.079) | 0.836 | -3.00 (0.003) |
| LDL-cholesterol | 0.161 (0.076) | 0.035 | -0.052 (0.074) | 0.480 | 2.01 (0.044) | 0.283 (0.085) | 0.001 | -0.087 (0.083) | 0.297 | 3.11 (0.002) |
| TG:HDL-c | 0.345 (0.073) | <0.001 | 0.004 (0.070) | 0.958 | 3.38 (<0.001) | 0.378 (0.078) | <0.001 | -0.088 (0.077) | 0.254 | 4.26 (<0.001) |
| Total leukocyte count | 0.372 (0.075) | <0.001 | -0.164 (0.073) | 0.026 | 5.11 (<0.001) | 0.272 (0.079) | <0.001 | -0.075 (0.079) | 0.341 | 3.10 (0.002) |
| C-reactive protein | 0.459 (0.070) | <0.001 | -0.122 (0.068) | 0.074 | 5.95 (<0.001) | 0.524 (0.076) | <0.001 | -0.196 (0.075) | 0.010 | 6.73 (<0.001) |

Values are β (SE) from multivariable linear regression models, additionally adjusted for birth weight. Diff. b/w coefficients = difference between the ASAT and VAT β-coefficients within each sex, tested using a Wald’s test (Z statistic, and p-value shown in parentheses). ASAT, abdominal subcutaneous adipose tissue; VAT, visceral adipose tissue; SE, standard error; BP, blood pressure. Leptin and Total Adiponectin were not assessed at the 24-year follow-up*.*

Supplementary Table S11: Associations of ASAT and VAT at 18-years with cardiometabolic markers at 18- and 24years with an additional adjustment for birth length

|  | Men | | | | | Women | | | | |
| --- | --- | --- | --- | --- | --- | --- | --- | --- | --- | --- |
|  | ASAT (18-years) | | VAT (18-years) | | Diff. b/w coefficients | ASAT (18-years) | | VAT (18-years) | | Diff. b/w coefficients |
| Outcome | β (SE) | p-value | β (SE) | p-value | Z (p-value) | β (SE) | p-value | β (SE) | p-value | Z (p-value) |
| Outcomes assessed at 18-years | | | | | | | | | | |
| Fasting glucose | 0.015 (0.067) | 0.819 | 0.114 (0.068) | 0.094 | -1.03 (0.303) | 0.033 (0.067) | 0.624 | 0.102 (0.067) | 0.126 | -0.74 (0.461) |
| 120-minute glucose | 0.124 (0.067) | 0.066 | 0.067 (0.067) | 0.316 | 0.60 (0.548) | 0.129 (0.070) | 0.069 | -0.086 (0.070) | 0.220 | 2.16 (0.031) |
| Insulinogenic Index | 0.048 (0.069) | 0.483 | -0.050 (0.068) | 0.463 | 1.02 (0.309) | -0.003 (0.072) | 0.963 | -0.047 (0.072) | 0.516 | 0.43 (0.671) |
| Matsuda Index | -0.457 (0.057) | <0.001 | -0.139 (0.057) | 0.016 | -3.95 (<0.001) | -0.173 (0.069) | 0.012 | -0.063 (0.068) | 0.352 | -1.14 (0.255) |
| Disposition Index | -0.313 (0.063) | <0.001 | -0.118 (0.063) | 0.061 | -2.19 (0.029) | -0.198 (0.071) | 0.006 | 0.007 (0.070) | 0.924 | -2.06 (0.040) |
| Systolic BP | 0.189 (0.065) | 0.004 | 0.197 (0.065) | 0.003 | -0.08 (0.934) | 0.317 (0.064) | <0.001 | 0.011 (0.064) | 0.863 | 3.40 (<0.001) |
| Diastolic BP | 0.262 (0.066) | <0.001 | 0.040 (0.066) | 0.540 | 2.38 (0.017) | 0.371 (0.063) | <0.001 | -0.017 (0.063) | 0.791 | 4.37 (<0.001) |
| Triglycerides | 0.311 (0.063) | <0.001 | 0.056 (0.064) | 0.383 | 2.85 (0.004) | 0.193 (0.066) | 0.004 | 0.053 (0.066) | 0.418 | 1.50 (0.135) |
| Total cholesterol | 0.294 (0.064) | <0.001 | 0.074 (0.064) | 0.249 | 2.43 (0.015) | 0.188 (0.066) | 0.005 | -0.036 (0.066) | 0.587 | 2.39 (0.017) |
| HDL-cholesterol | -0.070 (0.068) | 0.302 | -0.011 (0.068) | 0.866 | -0.61 (0.542) | -0.070 (0.066) | 0.292 | -0.162 (0.066) | 0.015 | 0.99 (0.323) |
| LDL-cholesterol | 0.385 (0.064) | <0.001 | 0.001 (0.064) | 0.982 | 4.25 (<0.001) | 0.218 (0.066) | 0.001 | 0.015 (0.066) | 0.818 | 2.18 (0.029) |
| TG:HDL-c | 0.279 (0.064) | <0.001 | 0.049 (0.065) | 0.446 | 2.52 (0.012) | 0.187 (0.065) | 0.004 | 0.119 (0.065) | 0.070 | 0.74 (0.458) |
| Leptin | 0.814 (0.036) | <0.001 | 0.062 (0.036) | 0.087 | 14.81 (<0.001) | 0.772 (0.044) | <0.001 | -0.032 (0.044) | 0.465 | 12.82 (<0.001) |
| Total Adiponectin | -0.303 (0.064) | <0.001 | -0.101 (0.064) | 0.113 | -2.23 (0.026) | -0.258 (0.066) | <0.001 | 0.012 (0.066) | 0.857 | -2.88 (0.004) |
| Total leukocyte count | 0.161 (0.066) | 0.015 | 0.052 (0.066) | 0.431 | 1.16 (0.245) | 0.170 (0.066) | 0.010 | 0.013 (0.066) | 0.845 | 1.70 (0.090) |
| C-reactive protein | 0.155 (0.068) | 0.024 | 0.029 (0.068) | 0.671 | 1.31 (0.190) | 0.177 (0.073) | 0.016 | -0.054 (0.072) | 0.455 | 2.25 (0.025) |
| Outcomes assessed at 24-years | | | | | | | | | | |
| Fasting glucose | 0.130 (0.077) | 0.095 | 0.069 (0.075) | 0.363 | 0.57 (0.571) | 0.129 (0.080) | 0.107 | 0.107 (0.080) | 0.182 | 0.20 (0.844) |
| 120-minute glucose | 0.138 (0.077) | 0.073 | 0.087 (0.075) | 0.247 | 0.48 (0.632) | 0.124 (0.083) | 0.136 | -0.088 (0.083) | 0.291 | 1.81 (0.071) |
| Insulinogenic Index | -0.148 (0.077) | 0.055 | -0.059 (0.075) | 0.428 | -0.83 (0.408) | -0.183 (0.082) | 0.026 | -0.029 (0.082) | 0.729 | -1.34 (0.182) |
| Matsuda Index | -0.494 (0.068) | <0.001 | -0.001 (0.066) | 0.986 | -5.18 (<0.001) | -0.369 (0.077) | <0.001 | 0.076 (0.077) | 0.325 | -4.11 (<0.001) |
| Disposition Index | -0.453 (0.069) | <0.001 | -0.028 (0.067) | 0.673 | -4.40 (<0.001) | -0.372 (0.076) | <0.001 | 0.052 (0.077) | 0.501 | -3.92 (<0.001) |
| Systolic BP | 0.224 (0.075) | 0.003 | 0.071 (0.073) | 0.333 | 1.46 (0.145) | 0.271 (0.079) | <0.001 | 0.009 (0.080) | 0.908 | 2.33 (0.020) |
| Diastolic BP | 0.284 (0.075) | <0.001 | 0.010 (0.073) | 0.889 | 2.62 (0.009) | 0.348 (0.079) | <0.001 | -0.107 (0.079) | 0.181 | 4.06 (<0.001) |
| Triglycerides | 0.331 (0.073) | <0.001 | 0.008 (0.071) | 0.915 | 3.18 (0.001) | 0.231 (0.081) | 0.005 | -0.130 (0.081) | 0.113 | 3.14 (0.002) |
| Total cholesterol | 0.214 (0.077) | 0.006 | -0.057 (0.075) | 0.449 | 2.51 (0.012) | 0.148 (0.083) | 0.077 | -0.107 (0.083) | 0.199 | 2.17 (0.030) |
| HDL-cholesterol | -0.197 (0.077) | 0.011 | 0.014 (0.075) | 0.855 | -1.97 (0.049) | -0.341 (0.078) | <0.001 | -0.029 (0.078) | 0.713 | -2.82 (0.005) |
| LDL-cholesterol | 0.191 (0.078) | 0.014 | -0.079 (0.076) | 0.298 | 2.49 (0.013) | 0.263 (0.082) | 0.002 | -0.085 (0.082) | 0.298 | 3.01 (0.003) |
| TG:HDL-c | 0.351 (0.072) | <0.001 | 0.002 (0.070) | 0.982 | 3.49 (<0.001) | 0.341 (0.079) | <0.001 | -0.095 (0.079) | 0.232 | 3.90 (<0.001) |
| Total leukocyte count | 0.359 (0.075) | <0.001 | -0.158 (0.073) | 0.030 | 4.96 (<0.001) | 0.242 (0.080) | 0.003 | -0.088 (0.081) | 0.279 | 2.89 (0.004) |
| C-reactive protein | 0.471 (0.071) | <0.001 | -0.133 (0.069) | 0.056 | 6.11 (<0.001) | 0.512 (0.073) | <0.001 | -0.205 (0.073) | 0.005 | 6.96 (<0.001) |

Values are β (SE) from multivariable linear regression models. Diff. b/w coefficients = difference between the ASAT and VAT β-coefficients within each sex, tested using a Wald’s test (Z statistic, and p-value shown in parentheses). ASAT, abdominal subcutaneous adipose tissue; VAT, visceral adipose tissue; SE, standard error; BP, blood pressure. Leptin and Total Adiponectin were not assessed at the 24-year follow-up.

Supplementary Table S12: Associations of ASAT and VAT at 18-years with cardiometabolic markers at 18- and 24years with an additional adjustment for birth head circumference

|  | Men | | | | | Women | | | | |
| --- | --- | --- | --- | --- | --- | --- | --- | --- | --- | --- |
|  | ASAT (18-years) | | VAT (18-years) | | Diff. b/w coefficients | ASAT (18-years) | | VAT (18-years) | | Diff. b/w coefficients |
| Outcome | β (SE) | p-value | β (SE) | p-value | Z (p-value) | β (SE) | p-value | β (SE) | p-value | Z (p-value) |
| Outcomes assessed at 18-years | | | | | | | | | | |
| Fasting glucose | 0.020 (0.067) | 0.762 | 0.116 (0.067) | 0.085 | -1.01 (0.314) | 0.029 (0.067) | 0.667 | 0.097 (0.067) | 0.151 | -0.71 (0.475) |
| 120-minute glucose | 0.127 (0.067) | 0.061 | 0.058 (0.067) | 0.383 | 0.72 (0.470) | 0.118 (0.072) | 0.101 | -0.106 (0.071) | 0.141 | 2.21 (0.027) |
| Insulinogenic Index | 0.044 (0.068) | 0.523 | -0.049 (0.068) | 0.466 | 0.97 (0.333) | 0.006 (0.073) | 0.934 | -0.027 (0.072) | 0.707 | 0.32 (0.745) |
| Matsuda Index | -0.465 (0.056) | <0.001 | -0.136 (0.056) | 0.016 | -4.12 (<0.001) | -0.173 (0.068) | 0.012 | -0.067 (0.068) | 0.323 | -1.10 (0.272) |
| Disposition Index | -0.320 (0.063) | <0.001 | -0.117 (0.062) | 0.061 | -2.30 (0.021) | -0.192 (0.072) | 0.008 | 0.018 (0.071) | 0.799 | -2.07 (0.038) |
| Systolic BP | 0.194 (0.065) | 0.003 | 0.193 (0.064) | 0.003 | 0.01 (0.989) | 0.321 (0.064) | <0.001 | 0.018 (0.064) | 0.783 | 3.37 (<0.001) |
| Diastolic BP | 0.262 (0.066) | <0.001 | 0.037 (0.066) | 0.575 | 2.42 (0.016) | 0.372 (0.063) | <0.001 | -0.015 (0.063) | 0.808 | 4.37 (<0.001) |
| Triglycerides | 0.308 (0.063) | <0.001 | 0.051 (0.063) | 0.417 | 2.86 (0.004) | 0.185 (0.066) | 0.005 | 0.043 (0.066) | 0.518 | 1.53 (0.126) |
| Total cholesterol | 0.292 (0.064) | <0.001 | 0.072 (0.064) | 0.260 | 2.43 (0.015) | 0.185 (0.066) | 0.006 | -0.041 (0.067) | 0.538 | 2.40 (0.016) |
| HDL-cholesterol | -0.074 (0.068) | 0.278 | -0.008 (0.068) | 0.905 | -0.68 (0.494) | -0.067 (0.066) | 0.306 | -0.159 (0.066) | 0.017 | 0.98 (0.328) |
| LDL-cholesterol | 0.391 (0.064) | <0.001 | -0.003 (0.063) | 0.968 | 4.37 (<0.001) | 0.215 (0.066) | 0.001 | 0.010 (0.066) | 0.875 | 2.20 (0.028) |
| TG:HDL-c | 0.278 (0.064) | <0.001 | 0.045 (0.064) | 0.490 | 2.56 (0.011) | 0.180 (0.065) | 0.006 | 0.109 (0.065) | 0.096 | 0.78 (0.437) |
| Leptin | 0.813 (0.036) | <0.001 | 0.055 (0.036) | 0.130 | 14.87 (<0.001) | 0.770 (0.044) | <0.001 | -0.035 (0.044) | 0.435 | 12.81 (<0.001) |
| Total Adiponectin | -0.304 (0.065) | <0.001 | -0.093 (0.064) | 0.144 | -2.33 (0.020) | -0.258 (0.066) | <0.001 | 0.011 (0.066) | 0.863 | -2.87 (0.004) |
| Total leukocyte count | 0.155 (0.066) | 0.019 | 0.049 (0.066) | 0.461 | 1.15 (0.250) | 0.172 (0.066) | 0.009 | 0.015 (0.066) | 0.814 | 1.69 (0.092) |
| C-reactive protein | 0.147 (0.068) | 0.032 | 0.026 (0.067) | 0.694 | 1.26 (0.207) | 0.174 (0.073) | 0.018 | -0.057 (0.073) | 0.437 | 2.24 (0.025) |
| Outcomes assessed at 24-years | | | | | | | | | | |
| Fasting glucose | 0.133 (0.077) | 0.086 | 0.069 (0.075) | 0.355 | 0.60 (0.550) | 0.130 (0.079) | 0.104 | 0.115 (0.080) | 0.151 | 0.14 (0.892) |
| 120-minute glucose | 0.132 (0.077) | 0.087 | 0.077 (0.074) | 0.296 | 0.51 (0.612) | 0.120 (0.083) | 0.149 | -0.100 (0.083) | 0.233 | 1.87 (0.061) |
| Insulinogenic Index | -0.142 (0.076) | 0.064 | -0.058 (0.074) | 0.435 | -0.79 (0.427) | -0.184 (0.081) | 0.026 | -0.030 (0.082) | 0.713 | -1.32 (0.185) |
| Matsuda Index | -0.493 (0.068) | <0.001 | -0.001 (0.066) | 0.984 | -5.18 (<0.001) | -0.372 (0.077) | <0.001 | 0.066 (0.077) | 0.393 | -4.04 (<0.001) |
| Disposition Index | -0.449 (0.069) | <0.001 | -0.028 (0.067) | 0.678 | -4.39 (<0.001) | -0.376 (0.076) | <0.001 | 0.044 (0.077) | 0.568 | -3.88 (<0.001) |
| Systolic BP | 0.224 (0.075) | 0.003 | 0.066 (0.073) | 0.369 | 1.51 (0.131) | 0.275 (0.079) | <0.001 | 0.024 (0.080) | 0.762 | 2.23 (0.026) |
| Diastolic BP | 0.280 (0.075) | <0.001 | 0.003 (0.072) | 0.967 | 2.65 (0.008) | 0.350 (0.079) | <0.001 | -0.102 (0.079) | 0.202 | 4.03 (<0.001) |
| Triglycerides | 0.328 (0.073) | <0.001 | -0.004 (0.071) | 0.952 | 3.27 (0.001) | 0.234 (0.081) | 0.004 | -0.121 (0.081) | 0.140 | 3.08 (0.002) |
| Total cholesterol | 0.217 (0.077) | 0.005 | -0.058 (0.075) | 0.437 | 2.57 (0.010) | 0.148 (0.083) | 0.078 | -0.109 (0.083) | 0.195 | 2.17 (0.030) |
| HDL-cholesterol | -0.196 (0.077) | 0.012 | 0.022 (0.074) | 0.765 | -2.04 (0.041) | -0.342 (0.078) | <0.001 | -0.035 (0.078) | 0.655 | -2.78 (0.005) |
| LDL-cholesterol | 0.197 (0.077) | 0.012 | -0.078 (0.075) | 0.301 | 2.55 (0.011) | 0.263 (0.082) | 0.002 | -0.086 (0.082) | 0.295 | 3.02 (0.003) |
| TG:HDL-c | 0.349 (0.072) | <0.001 | -0.012 (0.070) | 0.869 | 3.58 (<0.001) | 0.344 (0.079) | <0.001 | -0.085 (0.079) | 0.285 | 3.84 (<0.001) |
| Total leukocyte count | 0.348 (0.074) | <0.001 | -0.154 (0.071) | 0.032 | 4.90 (<0.001) | 0.241 (0.080) | 0.003 | -0.093 (0.081) | 0.255 | 2.92 (0.004) |
| C-reactive protein | 0.473 (0.071) | <0.001 | -0.137 (0.068) | 0.046 | 6.19 (<0.001) | 0.511 (0.073) | <0.001 | -0.209 (0.073) | 0.004 | 6.99 (<0.001) |

Values are β (SE) from multivariable linear regression models. Diff. b/w coefficients = difference between the ASAT and VAT β-coefficients within each sex, tested using a Wald’s test (Z statistic, and p-value shown in parentheses). ASAT, abdominal subcutaneous adipose tissue; VAT, visceral adipose tissue; SE, standard error; BP, blood pressure. Leptin and Total Adiponectin were not assessed at the 24-year follow-up

Supplementary Table S13: Comparison between low and high VAT:ASAT ratio groups classified as below and above-median ratio

| **Outcome** | **sex** | **N Low** | **N High** | **Low - Median (25^th^-75^th^)_** | **High - Median (25^th^-75^th^)** | **p-value** | **sex** | **N Low** | **N High** | **Low - Median (25^th^-75^th^)_** | **High - Median (25^th^-75^th^)** | **p-value** |
| --- | --- | --- | --- | --- | --- | --- | --- | --- | --- | --- | --- | --- |
| Birth weight | Women | 135 | 130 | 2.55 (2.30-2.80) | 2.50 (2.30-2.75) | 0.375 | Men | 144 | 146 | 2.75 (2.50-3.00) | 2.70 (2.46-2.94) | 0.204 |
| Age | Women | 140 | 141 | 17.61 (17.16-17.99) | 17.67 (17.23-18.10) | 0.318 | Men | 155 | 155 | 18.11 (17.86-18.53) | 18.20 (17.89-18.58) | 0.483 |
| BMI | Women | 140 | 141 | 19.76 (18.00-21.96) | 17.05 (15.80-18.46) | **<0.001** | Men | 155 | 155 | 21.11 (19.33-23.83) | 17.82 (16.53-18.88) | **<0.001** |
| Waist Circumference | Women | 140 | 141 | 71.12 (67.29-74.56) | 64.55 (61.85-68.00) | **<0.001** | Men | 155 | 155 | 76.20 (71.88-84.50) | 67.20 (64.00-70.38) | **<0.001** |
| Fasting glucose | Women | 140 | 141 | 93.00 (89.00-96.00) | 93.00 (89.00-96.00) | 0.684 | Men | 155 | 155 | 98.00 (94.00-100.00) | 97.00 (93.00-101.00) | 0.63 |
| 120-minute glucose | Women | 118 | 130 | 114.50 (101.25-130.00) | 114.00 (96.25-126.50) | 0.162 | Men | 153 | 153 | 114.00 (99.00-128.00) | 106.00 (94.00-124.00) | **0.011** |
| Insulinogenic Index | Women | 117 | 130 | 1.70 (1.42-2.10) | 1.68 (1.35-2.04) | 0.551 | Men | 153 | 148 | 1.72 (1.45-2.08) | 1.67 (1.33-2.07) | 0.26 |
| Matsuda Index | Women | 131 | 134 | 10.95 (8.28-15.40) | 12.25 (9.80-16.32) | **0.027** | Men | 154 | 153 | 11.37 (8.08-16.87) | 16.53 (12.44-20.84) | **<0.001** |
| Disposition Index | Women | 117 | 130 | 4.05 (3.64-4.49) | 4.18 (3.76-4.59) | 0.076 | Men | 153 | 149 | 4.19 (3.69-4.69) | 4.43 (4.01-4.98) | **<0.001** |
| Systolic BP | Women | 138 | 140 | 107.00 (100.00-111.75) | 104.00 (99.00-109.00) | 0.018 | Men | 155 | 152 | 114.00 (107.00-120.00) | 111.00 (104.00-117.25) | **0.021** |
| Diastolic BP | Women | 138 | 140 | 62.00 (58.00-66.00) | 59.50 (55.00-63.00) | **<0.001** | Men | 155 | 152 | 60.00 (54.00-65.00) | 56.00 (51.75-64.00) | **0.005** |
| Triglycerides | Women | 139 | 141 | 57.00 (46.00-68.50) | 51.00 (42.00-63.00) | **0.01** | Men | 155 | 153 | 69.00 (50.50-87.50) | 56.00 (45.00-67.00) | **<0.001** |
| Total cholesterol | Women | 139 | 141 | 137.00 (121.50-150.00) | 128.00 (114.00-142.00) | **0.002** | Men | 155 | 153 | 130.00 (115.00-147.00) | 117.00 (106.00-129.00) | **<0.001** |
| HDL-cholesterol | Women | 139 | 141 | 41.00 (36.00-48.00) | 42.00 (38.00-47.00) | 0.327 | Men | 155 | 153 | 36.00 (32.00-40.50) | 39.00 (34.00-43.00) | **0.027** |
| LDL-cholesterol | Women | 139 | 141 | 81.40 (69.30-93.60) | 73.00 (60.60-86.00) | **<0.001** | Men | 154 | 153 | 78.50 (64.65-89.95) | 66.80 (56.80-76.60) | **<0.001** |
| TG:HDL-c | Women | 139 | 141 | 1.36 (1.09-1.78) | 1.17 (0.98-1.51) | **0.01** | Men | 155 | 153 | 1.85 (1.26-2.65) | 1.44 (1.14-1.81) | **<0.001** |
| Leptin | Women | 140 | 140 | 23.25 (15.52-32.60) | 11.50 (8.17-16.52) | **<0.001** | Men | 155 | 154 | 6.50 (3.05-11.40) | 1.40 (0.78-2.50) | **<0.001** |
| Total Adiponectin | Women | 138 | 138 | 4.80 (3.90-6.06) | 5.80 (4.40-7.10) | **<0.001** | Men | 150 | 151 | 4.30 (3.42-5.60) | 5.00 (4.10-6.20) | **<0.001** |
| Total leukocyte count | Women | 140 | 140 | 6.70 (5.80-7.80) | 6.10 (5.20-7.12) | **0.008** | Men | 155 | 155 | 5.90 (5.10-6.80) | 5.70 (4.90-6.45) | 0.07 |
| C-reactive protein | Women | 116 | 122 | 0.08 (0.06-0.11) | 0.07 (0.04-0.10) | **0.038** | Men | 146 | 147 | 0.09 (0.06-0.15) | 0.08 (0.06-0.13) | 0.144 |

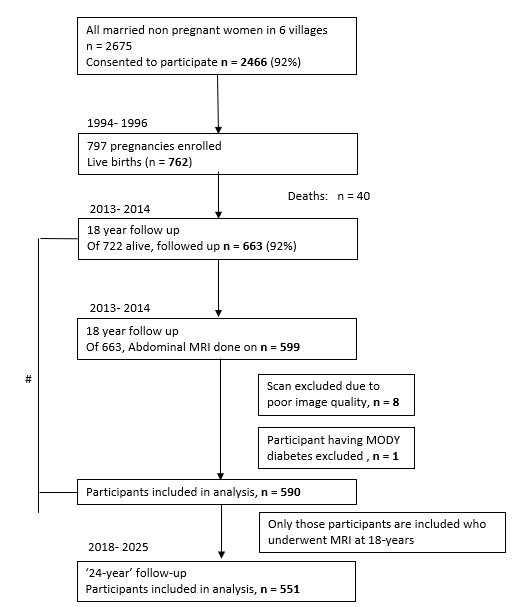

**Supplementary Figure S1:** **STROBE flow diagram of Pune Maternal Nutrition Study.** # 590 participants with MRI measurements were similar to 73 who did not have MRI, except for age and 120 min OGTT glucose.

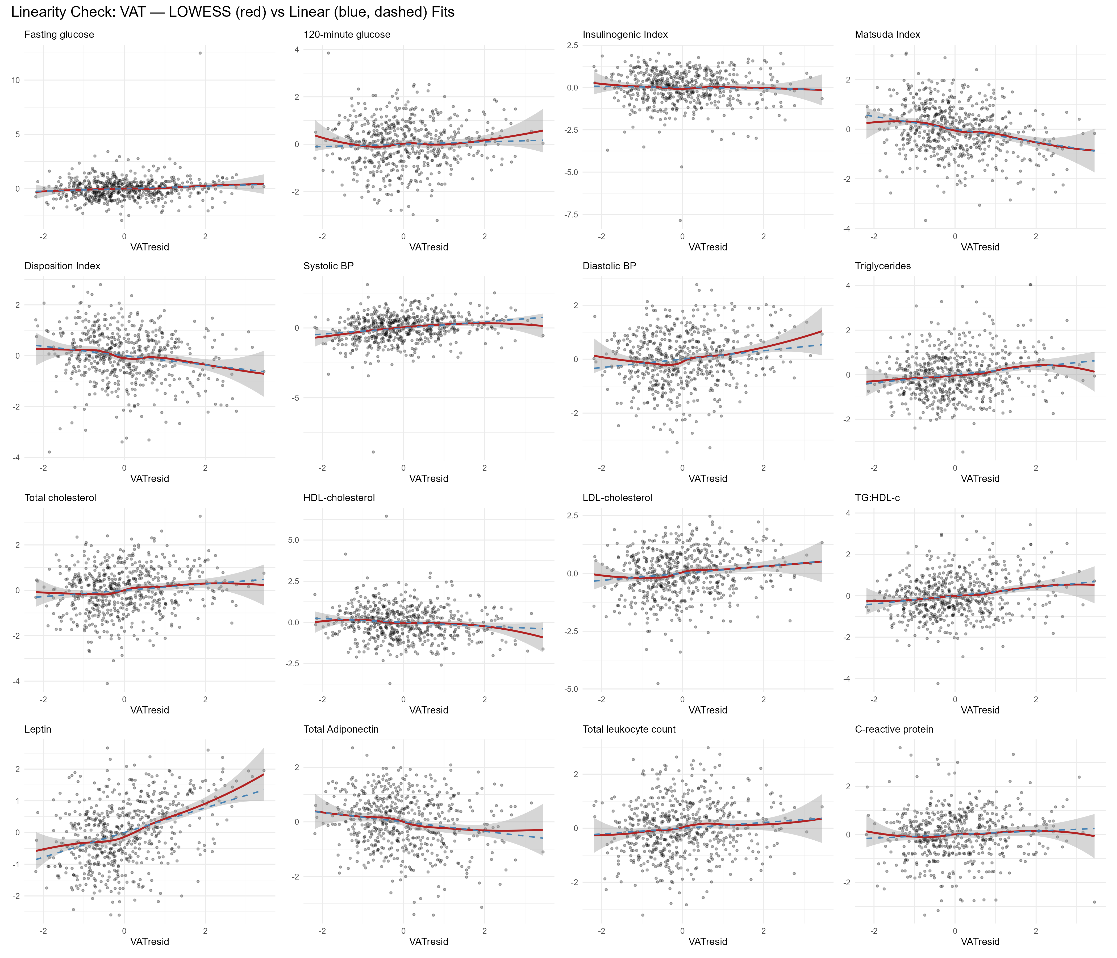

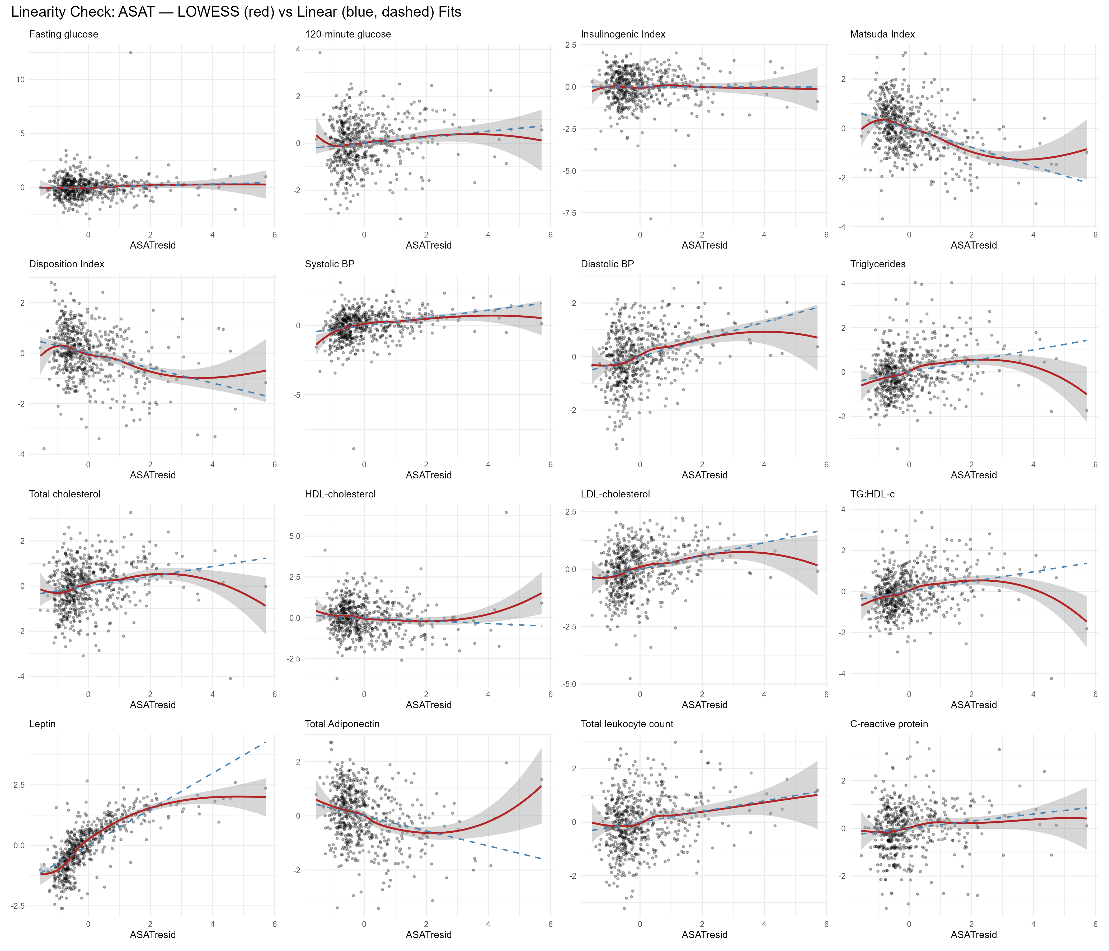

Supplementary Figure S2: LOWESS plots assessing linearity between outcome-exposure pairs (standardized residuals adjusted for age and socioeconomic status)

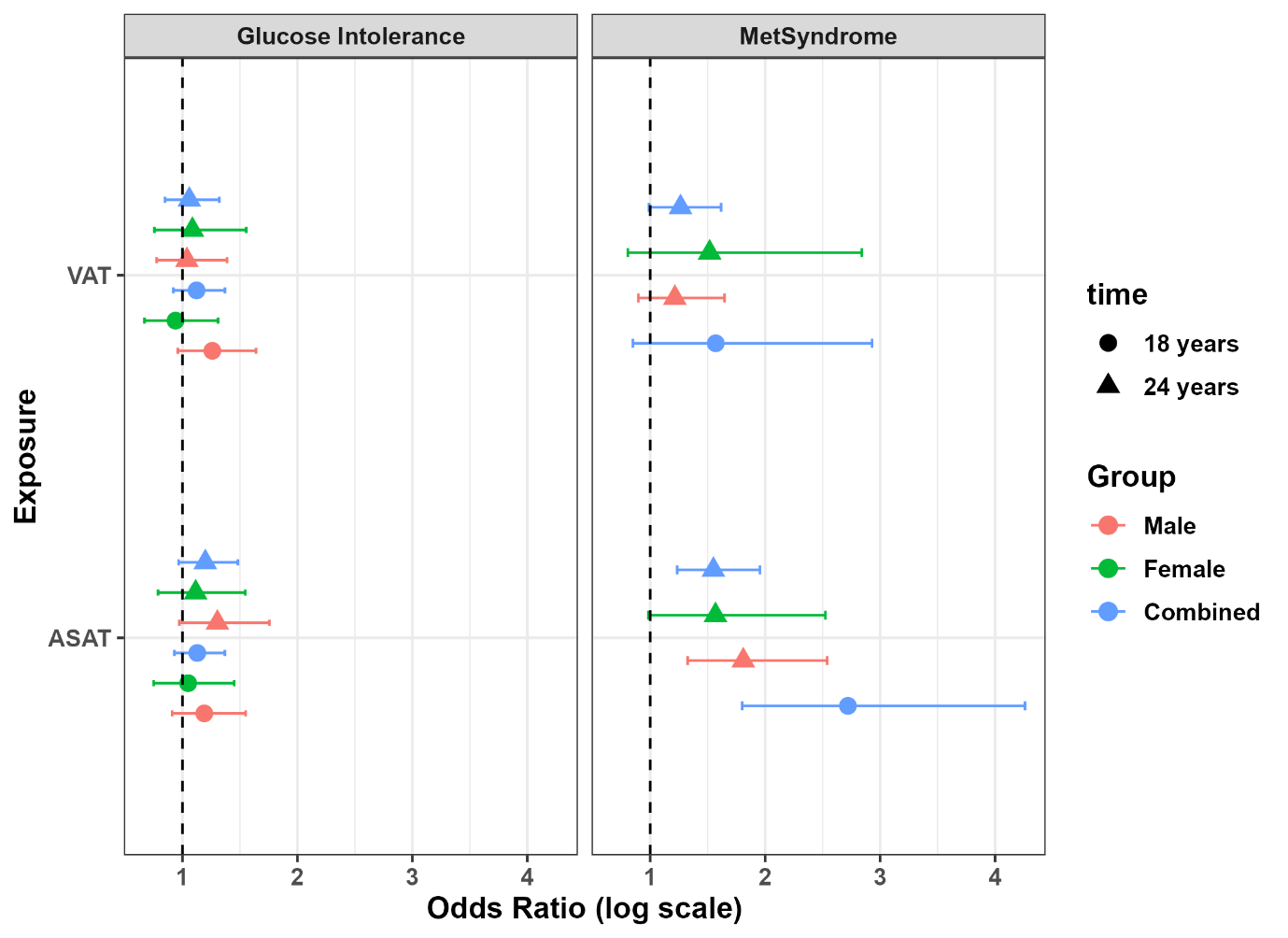

**Supplementary Figure S3**. Odds ratios for glucose intolerance at 18-years (124/310 Men and 54/280 Women) as well as 24-years (90/285 Men and 45/257 Women); and metabolic syndrome at 18-years (12/590) as well as 24-years (44/287 Men and 13/256 Women) as outcomes respectively. Given the small numbers of metabolic syndrome at 18-years, sex stratified analysis was not performed.
